# Effect of a single dose of 8 mg moxidectin or 150 µg/kg ivermectin on *O. volvulus* skin microfilariae in a randomized trial: Differences between areas in the Democratic Republic of the Congo, Liberia and Ghana and impact of intensity of infection

**DOI:** 10.1101/2022.02.22.22271335

**Authors:** Didier Bakajika, Eric M Kanza, Nicholas O Opoku, Hayford M Howard, Germain L Mambandu, Amos Nyathirombo, Maurice M Nigo, Kambale Kasonia Kennedy, Safari L Masembe, Mupenzi Mumbere, Kambale Kataliko, Kpehe M Bolay, Simon K Attah, George Olipoh, Sampson Asare, Michel Vaillant, Christine M Halleux, Annette C Kuesel

## Abstract

**Background:** Community-directed treatment with 150 µg/kg ivermectin (CDTI), the current control strategy, is insufficient for eliminating *Onchocerca volvulus* transmission across sub-Saharan Africa. Our study in CDTI-naïve areas in Nord Kivu and Ituri (Democratic Republic of the Congo (DRC), Lofa County (Liberia) and Nkwanta district (Ghana) showed that a single 8 mg moxidectin dose reduced skin microfilariae density (microfilariae/mg skin, SmfD) better and for longer than a single 150µg/kg ivermectin dose.

**Methodology/Principal Findings:** We analysed drug efficacy by study area and pre-treatment SmfD (intensity of infection, IoI). Four and three IoI categories were defined for across-study and by-study area analyses, respectively. We used a general linear model to analyse SmfD 1, 6, 12 and 18 months post-treatment, a logistic model to determine the odds of undetectable SmfD from month 1 to month 6 (UD1-6), month 12 (UD1-12) and month 18 (UD1-18), and descriptive statistics to quantitate inter-interindividual response differences. Twelve months post-treatment, treatment differences were 92.9%, 90.1%, 86.8% and 84.5% in Nord Kivu, Ituri, Lofa and Nkwanta, and 74.1%, 84.2%, 90.0% and 95.4% for participants with SmfD 10-20, ≥20-<50, ≥50-<80, ≥80, respectively. Ivermectin’s efficacy was lower in Ituri and Nkwanta than Nord Kivu and Lofa (p≤0.002) and moxidectin’s efficacy lower in Nkwanta than Nord Kivu, Ituri and Lofa (p<0.006). Odds ratios for UD1-6, UD1-12 or UD1-18 after moxidectin versus ivermectin treatment exceeded 7.0. Suboptimal response (SmfD 12 months post-treatment >40% of pre-treatment SmfD) occurred in 0%, 0.3%, 1.6% and 3.9% of moxidectin and 12.1%, 23.7%, 10.8% and 28.0% of ivermectin treated participants in Nord Kivu, Ituri, Lofa and Nkwanta, respectively.

**Conclusions/significance:** The benefit of moxidectin vs ivermectin treatment increased with pre-treatment IoI. The possibility that parasite populations in different areas have different drug susceptibility without prior ivermectin selection pressure needs to be considered and further investigated.

**Author Summary:** Onchocerciasis or river blindness is a parasitic disease primarily in sub-Saharan Africa and Yemen. It can cause debilitating morbidity including severe itching, skin changes, visual impairment and even blindness. Many years of control efforts, today primarily based on mass administration of ivermectin (MDA) in endemic communities, have reduced morbidity and the percentage of infected individuals so that elimination of parasite transmission is now planned. WHO estimated that in 2020 more than 239 million people required MDA. Ivermectin may not be sufficiently efficacious to achieve elimination everywhere. Our study in areas in Liberia, Ghana and the Democratic Republic of the Congo where MDA had not been implemented yet showed that moxidectin reduced parasite levels in the skin better and for longer than ivermectin. Here we show that people with higher numbers of parasites in the skin benefit more from moxidectin treatment than those with lower numbers and that the efficacy of ivermectin and moxidectin differs between study areas. Provided WHO and countries include moxidectin in guidelines and policies, this information could help decisions on when and where to use moxidectin.

## Introduction

Onchocerciasis is a vector-borne disease, caused by the helminth *Onchocerca volvulus*, endemic primarily in remote rural areas of sub-Saharan Africa. Different parasite life stages live in humans: the infective larvae transmitted by blackflies of the genus *Simulium* develop into macrofilariae which live primarily in subcutaneous and deep tissue nodules and produce for around 12 years millions of microfilariae which have an estimated life span of 1-2 years. The dermatological, lymphatic and ocular symptoms are due to the host inflammatory reactions to the dead microfilariae [1]. Evidence is increasing that *O. volvulus* infection is a risk factor for development of seizures and epilepsy [2].

Onchocerciasis morbidity, including severe itching, visual impairment and blindness, and its socio-economic impact have motivated large scale control programmes. Initially implemented in West Africa using vector control, these programmes are now based on mass drug administration (MDA) of ivermectin to the ’eligible population’ (≥5 years or ≥90 cm, not pregnant or acutely sick) [1]. In the six Central and South American countries with an estimated 0.56 million people living in areas where the parasite was transmitted, between 23 and 36 rounds of biannual ivermectin MDA, complemented with quarterly MDA in around 300 communities, have eliminated onchocerciasis [3]. The exception is the endemic area across the border between Venezuela and Brazil where around 35,000 highly mobile people required MDA in 2020 [4, 5]. In Sudan and Yemen, 0.79 million people required MDA in 2020 [5]. Onchocerciasis elimination faces the biggest challenge in Africa where >100 Million people live in meso- or hyperendemic areas [6–8] and more than 239 million were estimated to require MDA in 2020 [5]. In 11 countries, the Onchocerciasis Control Programme of West Africa (OCP, 1974-2002) eliminated onchocerciasis as a public health problem in the savannah areas through large scale weekly aerial larviciding of vector breeding sites along around 50,000 km of rivers, complemented later by ivermectin MDA [1]. In the other countries, the African Programme for Onchocerciasis (APOC, 1995-2015 [9]) facilitated ivermectin MDA implemented via community directed treatment with ivermectin (CDTI) in meso- and hyperendemic areas. Long term CDTI has significantly decreased morbidity [10, 11]. For many years, it was, however, considered impossible to eliminate parasite transmission across Africa with CDTI [12]. This has motivated investments into discovery and development of more effective anti-onchocercal drugs [13]. A research study and APOC-supported evaluations of infection prevalence in areas under long term CDTI suggested that CDTI may be more effective than initially expected and could eliminate parasite transmission in many areas [14–17]. This resulted in the objective to eliminate onchocerciasis transmission in some African countries by 2020 and in 80% of endemic countries by 2025 [18, 19]. These targets have recently been revised to achieve WHO- verified interruption of parasite transmission in 12 countries world-wide by 2030 [20].

Expert consultations and detailed analyses of available data spearheaded by APOC concluded that alternative treatment strategies (ATS), including those with more effective drugs than ivermectin, are needed to eliminate parasite transmission in many areas [21].

The development of moxidectin, a milbemycin macrocyclic lactone, for onchocerciasis was initiated by the UNICEF/UNDP/World Bank/WHO Special Programme for Research and Training in Tropical Diseases (TDR) through pre-clinical pharmacology studies. Their results, combined with the non-clinical toxicology data acquired for registration of moxidectin for veterinary use (Document 2 in Annex 2 in [22], identified moxidectin as a clinical development candidate. Six Phase 1 studies in healthy volunteers were conducted [23–28]. A Phase 2 and a Phase 3 study in *O. volvulus* infected individuals [29, 30] showed that a single oral dose of 8 mg moxidectin reduces skin microfilariae (mf) density (microfilariae/mg skin, SmfD) better and prevents repopulation of the skin with microfilariae for longer than a single oral dose of ivermectin as used during CDTI (150 µg/kg). In the Phase 3 study, the treatment difference (calculated as the difference between the adjusted geometric mean SmfD in the two treatment arms in percentage of the adjusted geometric mean SmfD in the ivermectin treatment arm) was 86%, 97%, 86% and 76% at 1, 6, 12 and 18 months after treatment (p<0.0001). The treatment difference was independent of sex, but higher for individuals with ≥20 SmfD (93% at month 12) than for individuals with <20 SmfD (76.0% at month 12) pre-treatment [30]. Both studies showed that moxidectin and ivermectin have comparable safety profiles. In conjunction with the safety data from the healthy volunteer studies and non-clinical toxicology studies, the Phase 2 and Phase 3 study safety data suggest that moxidectin may have the safety profile required for Community-Directed Treatment with Moxidectin (CDTM). The Food and Drug Administration of the United States of America (US FDA) approved moxidectin for the treatment of onchocerciasis in ≥12-year-olds on 13 June 2018 following priority review of a New Drug Application submitted by Medicines Development for Global Health (MDGH). MDGH is the Australian not-for-profit biopharmaceutical company to whom WHO had licensed all moxidectin- related data at its disposal to register moxidectin. [31]

The previous report of the Phase 3 study [30] analyzed the efficacy of moxidectin and ivermectin across all participants and by the two pre-treatment SmfD categories used for stratifying participants at randomization. Here, we are reporting analyses conducted to investigate whether the efficacy of the drugs differs between study areas, to better characterize the dependency of the treatment difference on pre-treatment SmfD and to explore the significant inter-individual variability of response to ivermectin the raw data suggested [30]. Furthermore, we are reporting the *O. volvulus* infection relevant data obtained in all individuals screened by study area.

## Methods

Details for all methods except the statistical analyses for this manuscript have been provided previously [30] and are only summarized briefly.

### Trial registration

The study was registered on 14 November 2008 in Clinicaltrials.gov (ID: NCT00790998).

### Ethics Committee approval and participant consent

Conduct of this study, including protocol, information documents for potential participants and the participant consent forms, were approved by the Ghana Food and Drugs Authority and the Ghana Health Service Ethics Review Committee, the Liberia Ministry of Health and Social Welfare and the Ethics Committee of the Liberia Institute for Biomedical Research, the Ministère de la Santé Publique of DRC and the Ethics Committee of the Ecole de la Santé Publique Université de Kinshasa in DRC, and the WHO Ethics Review Committee.

Volunteers provided their consent or assent with parental consent to study participation through signature or thumbprint in the presence of a literate witness in their villages. This included consent to publication of summaries of the results.

### Study design and objectives

The double-blind, randomized, ivermectin-controlled study in *O. volvulus* infected individuals was designed to determine whether a single 8 mg oral dose of moxidectin achieves SmfD ≤50% of SmfD after a single oral dose of 150µg/kg ivermectin one year after treatment and to collect safety data.

### Study timing and areas

The study was conducted from 2009 to 2012 in four onchocerciasis endemic areas in which CDTI had not yet been initiated: Nord Kivu province (current Zones de Santé Kalunguta and

Mabalako) in the Democratic Republic of the Congo (DRC), Ituri province in DRC (Zone de Santé Logo in Northern Ituri, subsequently referred to as Nord Ituri), Lofa county in Liberia (subsequently referred to as Lofa or Liberia) and the Kpasa subdistrict within the Nkwanta North health district in Ghana (subsequently referred to as Nkwanta or Ghana).

### Participant eligibility criteria

Volunteers had to be ≥12 years old, weigh ≥30 kg and have ≥10 SmfD. Women of reproductive capacity could not be pregnant and had to commit to using reliable contraception (abstinence, condoms or hormonal contraception are medically recognized methods) for 6 months after treatment (for ivermectin and moxidectin labelling regarding use in pregnant women see, respectively, https://www.merck.com/product/usa/pi_circu/s/stromectol/strmectol_pi.pdf and https://www.accessd.fda.gov/ipts/cder/daf/index.cfm?event=overview.process&varApplNo=210867). To include a population as similar to that which would be participating in CDTM as possible, volunteers meeting inclusion criteria consistent with available pre-clinical and clinical safety data were excluded only if they met criteria potentially compromising their safety, efficacy assessments or safety assessments.

### Randomization and treatment

Eligible participants were randomized by study area, sex and pre-treatment SmfD (<20 vs. ≥ 20 mf/mg skin) in a ratio of 2:1 to 8 mg moxidectin or 150 µg/kg ivermectin using computer-generated randomization lists. To ensure double blinding, a pharmacist not involved in participant evaluation provided for each participant four identical looking capsules containing 2 mg moxidectin tables, 3 mg ivermectin tablets or placebo, as required by treatment allocation and weight. A study team member observed participants while they were swallowing the capsules.

### Measurement of skin microfilariae densities

Four skin snips were obtained (one snip from each iliac crest and calf) pre-treatment and 1, 6, 12 and 18 months post-treatment and SmfD (microfilariae/mg skin) determined as described previously [29, 30].

### Duration of follow up

The study was initiated with last follow up occurring 18 months post-treatment. From July 2011 onward, WHO was the sole study sponsor. Resulting resource limitations required a protocol amendment eliminating the 18 months follow up examinations. This resulted in 96% of moxidectin treated and 97% of ivermectin treated participants with 12 months follow up but only 78% of participants in both treatment arms with month 18 follow up data [30].

### Sample size

The sample size required to show a ≥50% difference in the primary efficacy outcome SmfD 12 month after treatment with α=0.05 and 90% power and the 2:1 (moxidectin:ivermectin) randomization ratio was 185. For a better characterization of the safety profile, the planned sample size was 1000 moxidectin and 500 ivermectin treated individuals.

### Statistical analysis for this manuscript

In analogy to the Community Microfilarial Load [32], the microfilarial load among screened individuals (ScMFL) was calculated as the geometric mean of (SmfD+1) -1 for all villages in whom at least 18 individuals ≥20 years old were screened.

For analysis by pre-treatment SmfD, four SmfD categories, referred to as ‘intensity of infection’ (IoI) categories, were defined for analyses across study areas: 10 to <20, 20 to < 50, 50 to < 80 and ≥ 80. Considering the number of participants with ≥80 pre-treatment SmfD, only three IoI categories (10 to <20, 20 to < 50, ≥ 50) were defined for analyses by study area.

For inferential statistical analysis, SmfD were log-transformed (y=log_e_(SmfD+1)) before analysis.

Longitudinal analysis of SmfD 1, 6, 12 and 18 months after treatment used a general linear model for repeated measures with pre-treatment SmfD, treatment, sex, IoI category, time, treatment*IoI category, and treatment*time interaction as fixed effects, time as repeated effect for the four SmfD measurements with participant as the statistical unit. In the across study area analysis, study area was a random effect (for sensitivity analysis regarding study area as fixed or random effect see [30]). The treatment difference was calculated as the difference in adjusted geometric means of the two treatment arms in percentage of the adjusted geometric means in the ivermectin arm. Pairwise comparison of the efficacy of ivermectin and of moxidectin between the four study areas was conducted through marginal means comparisons (least squares means from the model). All participants who received study drug were included in these analyses (modified Intent to Treat population, mITT).

The odds of participants having undetectable SmfD at month 1 and 6 (UD1-6), at month 1, 6 and 12 (UD1-12) and at month 1, 6, 12 and 18 (UD1-18) after treatment with moxidectin or ivermectin were calculated using a logistic model with treatment, lol category and IoI category*treatment interaction as fixed effects. For the across study area analysis, the study area was a random effect. Only individuals who had SmfD determined at each of the relevant time points were included in these analyses. Note that the statistical analysis plan signed off prior to unblinding planned on analysis of percentage of participants with undetectable SmfD by measurement time point as presented in [30]. The analysis of UD 1-6, UD1-12 and UD1-18 was added.

Inter-individual differences in response to ivermectin and moxidectin between and within study areas and the extent to which the response might be explained by the pre-treatment SmfD, was explored with descriptive analyses by study area and IoI category.

Descriptive statistics and figures were generated with STATA 13.1 (StataCorp LLC, USA) or Excel 2016. Inferential statistical analyses were conducted with SAS System version 9.4 (SAS Institute, Cary, NC, USA).

## Results

### Consort diagram

The CONSORT flow diagram has been reported previously [30].

### Prevalence and intensity of *O. volvulus* infection among screened individuals

Table 1 shows for each study area the number of individuals screened by sex, age group and whether skin microfilariae were detected (0 mf/mg skin vs. >0 mf/mg skin). Presentation by <20 and ≥20 years was chosen because evaluation of 30-50 men ≥ 20 years for infection as indicated by the presence of palpable subcutaneous nodules is the basis for rapid epidemiological assessment (REA) and rapid epidemiological mapping of onchocerciasis prevalence (REMO). REA and REMO were used to identify and map onchocerciasis endemicity in the APOC countries. Percentages of ≥20 year old individuals with palpable nodules of approximately 20-30% and above approximately 30%, respectively, indicate meso-endemic (infection prevalence ≈40-60 %) and hyperendemic areas (infection prevalence above ≈60%) [33–35]. In these areas CDTI was to be implemented to eliminate onchocerciasis as a public health problem [6,7,9,36,37] since infection prevalence >40% was found to be associated with an increased risk of onchocercal blindness [38].

**Table 1:**
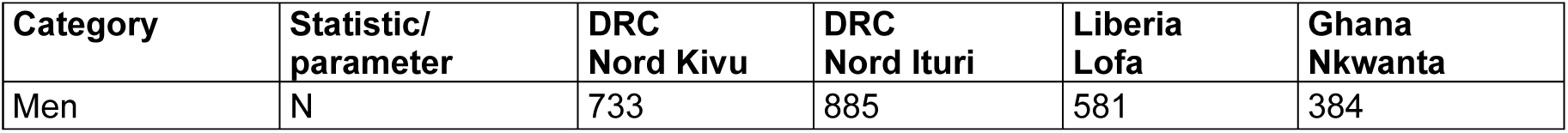
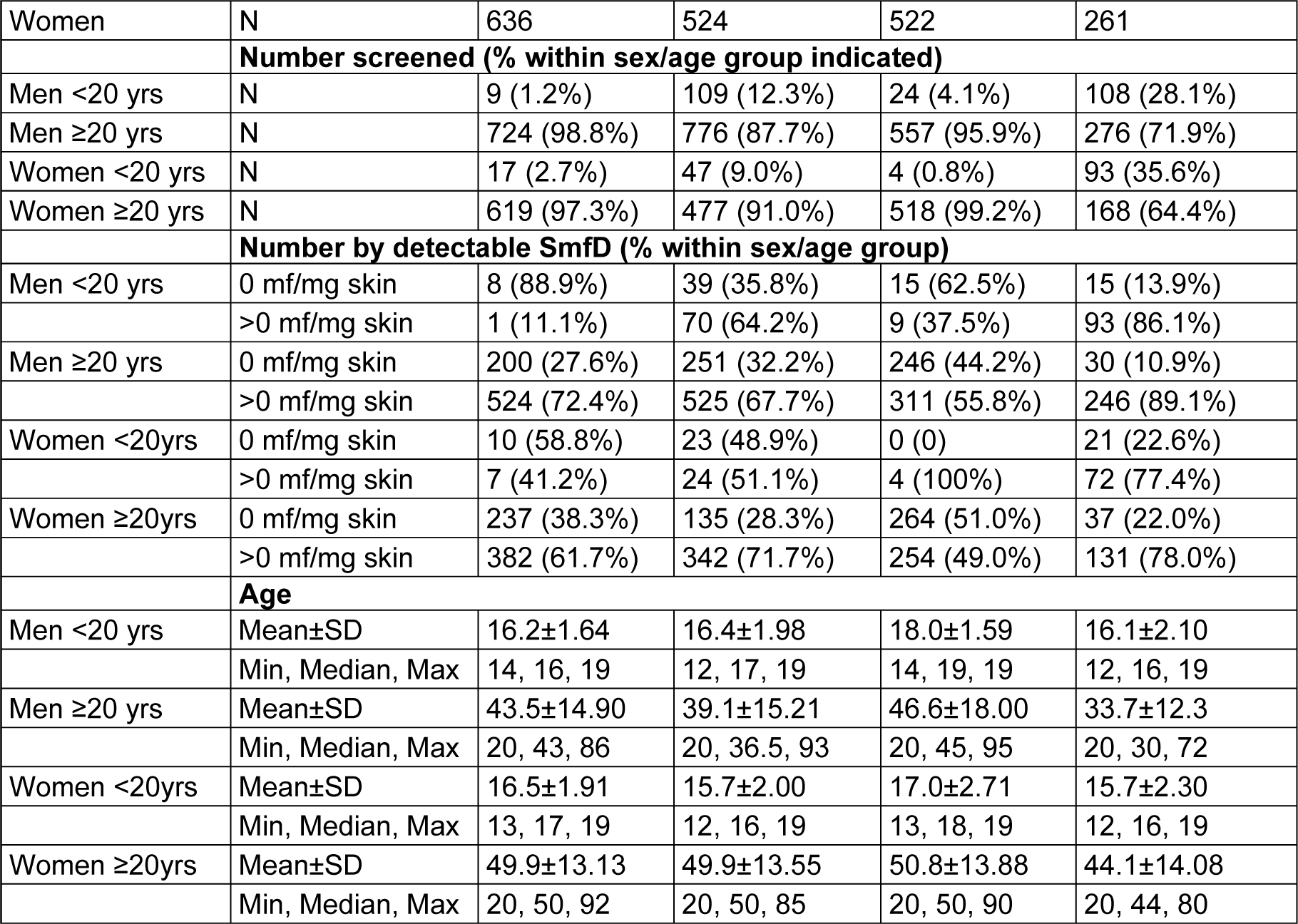
Sex, age and O. volvulus infection of individuals screened by study area and village.

| Category | Statistic/<br>parameter | DRC<br>Nord Kivu | DRC<br>Nord Ituri | Liberia<br>Lofa | Ghana<br>Nkwanta |
| --- | --- | --- | --- | --- | --- |
| Men | N | 733 | 885 | 581 | 384 |
| Women | N | 636 | 524 | 522 | 261 |
| **Number screened (% within sex/age group indicated)** |  |  |  |  |  |
| Men <20 yrs | N | 9 (1.2%) | 109 (12.3%) | 24 (4.1%) | 108 (28.1%) |
| Men ≥20 yrs | N | 724 (98.8%) | 776 (87.7%) | 557 (95.9%) | 276 (71.9%) |
| Women <20 yrs | N | 17 (2.7%) | 47 (9.0%) | 4 (0.8%) | 93 (35.6%) |
| Women ≥20 yrs | N | 619 (97.3%) | 477 (91.0%) | 518 (99.2%) | 168 (64.4%) |
| **Number by detectable SmfD (% within sex/age group)** |  |  |  |  |  |
| Men <20 yrs | 0 mf/mg skin | 8 (88.9%) | 39 (35.8%) | 15 (62.5%) | 15 (13.9%) |
|  | >0 mf/mg skin | 1 (11.1%) | 70 (64.2%) | 9 (37.5%) | 93 (86.1%) |
| Men ≥20 yrs | 0 mf/mg skin | 200 (27.6%) | 251 (32.2%) | 246 (44.2%) | 30 (10.9%) |
|  | >0 mf/mg skin | 524 (72.4%) | 525 (67.7%) | 311 (55.8%) | 246 (89.1%) |
| Women <20yrs | 0 mf/mg skin | 10 (58.8%) | 23 (48.9%) | 0 (0) | 21 (22.6%) |
|  | >0 mf/mg skin | 7 (41.2%) | 24 (51.1%) | 4 (100%) | 72 (77.4%) |
| Women ≥20yrs | 0 mf/mg skin | 237 (38.3%) | 135 (28.3%) | 264 (51.0%) | 37 (22.0%) |
|  | >0 mf/mg skin | 382 (61.7%) | 342 (71.7%) | 254 (49.0%) | 131 (78.0%) |
| **Age** |  |  |  |  |  |
| Men <20 yrs | Mean±SD | 16.2±1.64 | 16.4±1.98 | 18.0±1.59 | 16.1±2.10 |
|  | Min, Median, Max | 14, 16, 19 | 12, 17, 19 | 14, 19, 19 | 12, 16, 19 |
| Men ≥20 yrs | Mean±SD | 43.5±14.90 | 39.1±15.21 | 46.6±18.00 | 33.7±12.3 |
|  | Min, Median, Max | 20, 43, 86 | 20, 36.5, 93 | 20, 45, 95 | 20, 30, 72 |
| Women <20yrs | Mean±SD | 16.5±1.91 | 15.7±2.00 | 17.0±2.71 | 15.7±2.30 |
|  | Min, Median, Max | 13, 17, 19 | 12, 16, 19 | 13, 18, 19 | 12, 16, 19 |
| Women ≥20yrs | Mean±SD | 49.9±13.13 | 49.9±13.55 | 50.8±13.88 | 44.1±14.08 |
|  | Min, Median, Max | 20, 50, 92 | 20, 50, 85 | 20, 50, 90 | 20, 44, 80 |

Fig 1, Fig 2 and Fig 3 provide overviews of SmfD among those screened by study area. Even taking into account that (a) no consideration was given to choosing villages for recruitment based on being first or higher line villages and that (b) those volunteering for screening were not randomly selected and may be biased towards those suspecting themselves to be infected, the screening data show that in each study area, onchocerciasis was meso- or hyperendemic.

**Fig 1:**
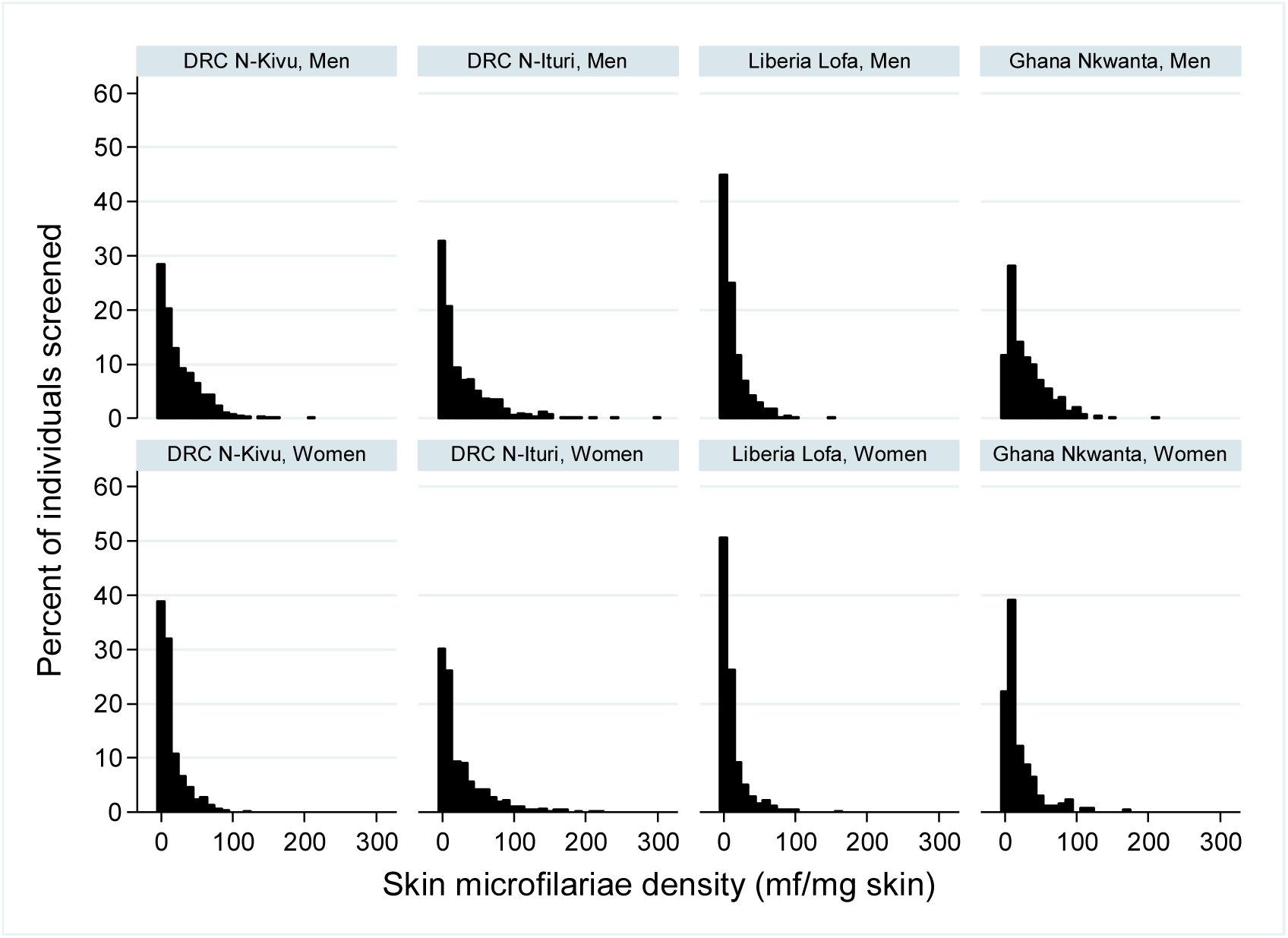
Distribution of skin microfilariae density among those screened by study area and sex.

**Fig 2:**
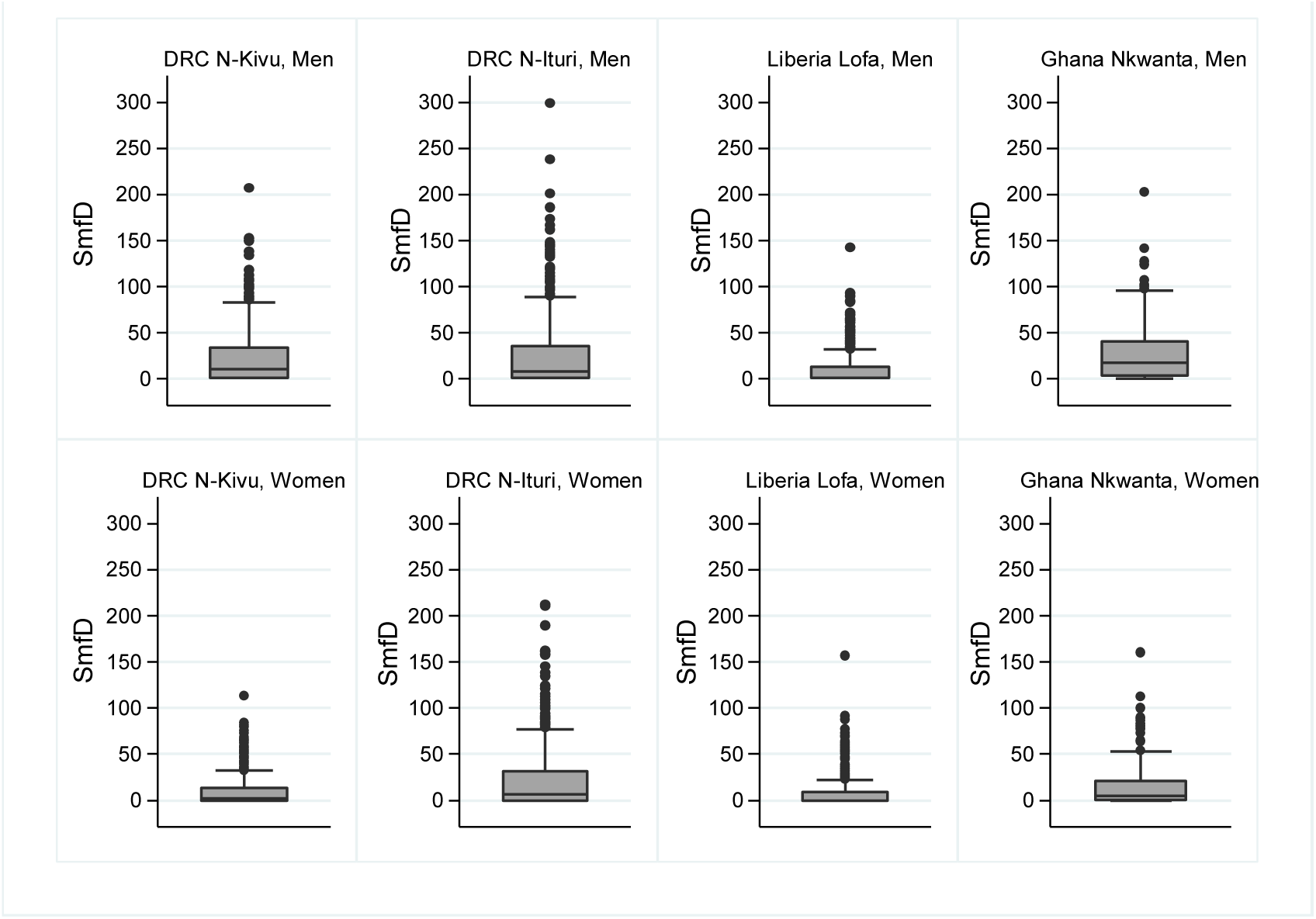
Skin microfilariae density across all individuals screened by study area and sex.

**Fig 3:**
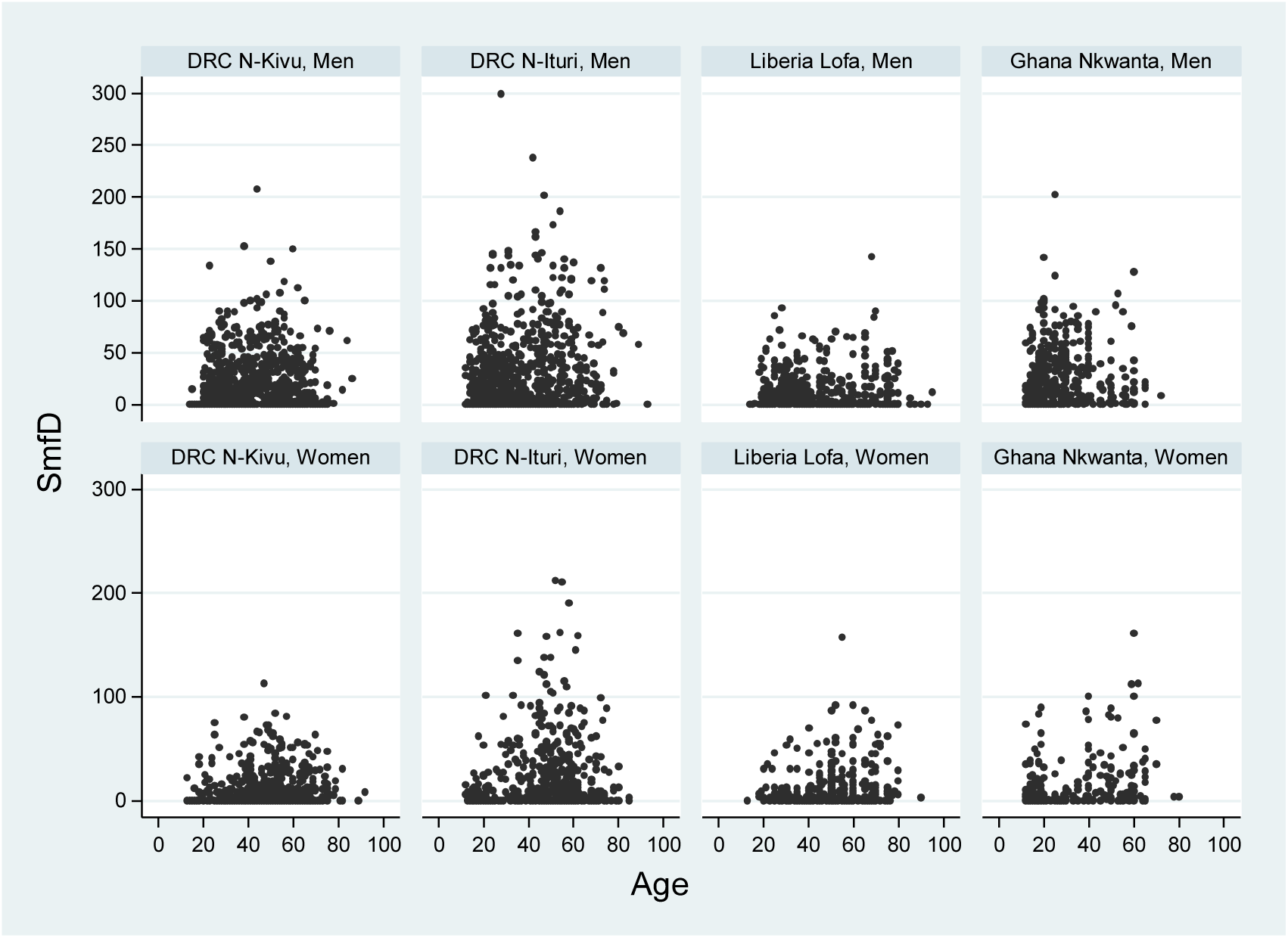
Skin microfilariae density among those screened by study area, sex and age.

Table 2 shows for villages where at least 18 individuals were screened two indicators of onchocerciasis endemicity: the percentage of ≥20 year olds with detectable skin microfilariae levels and their screening microfilarial load (ScMFL), calculated in the same way as the Community Microfilarial Load (CMFL) [32]. GPS coordinates of the villages from where the majority of participants were recruited are provided in S1 Table S1.

**Table 2:**
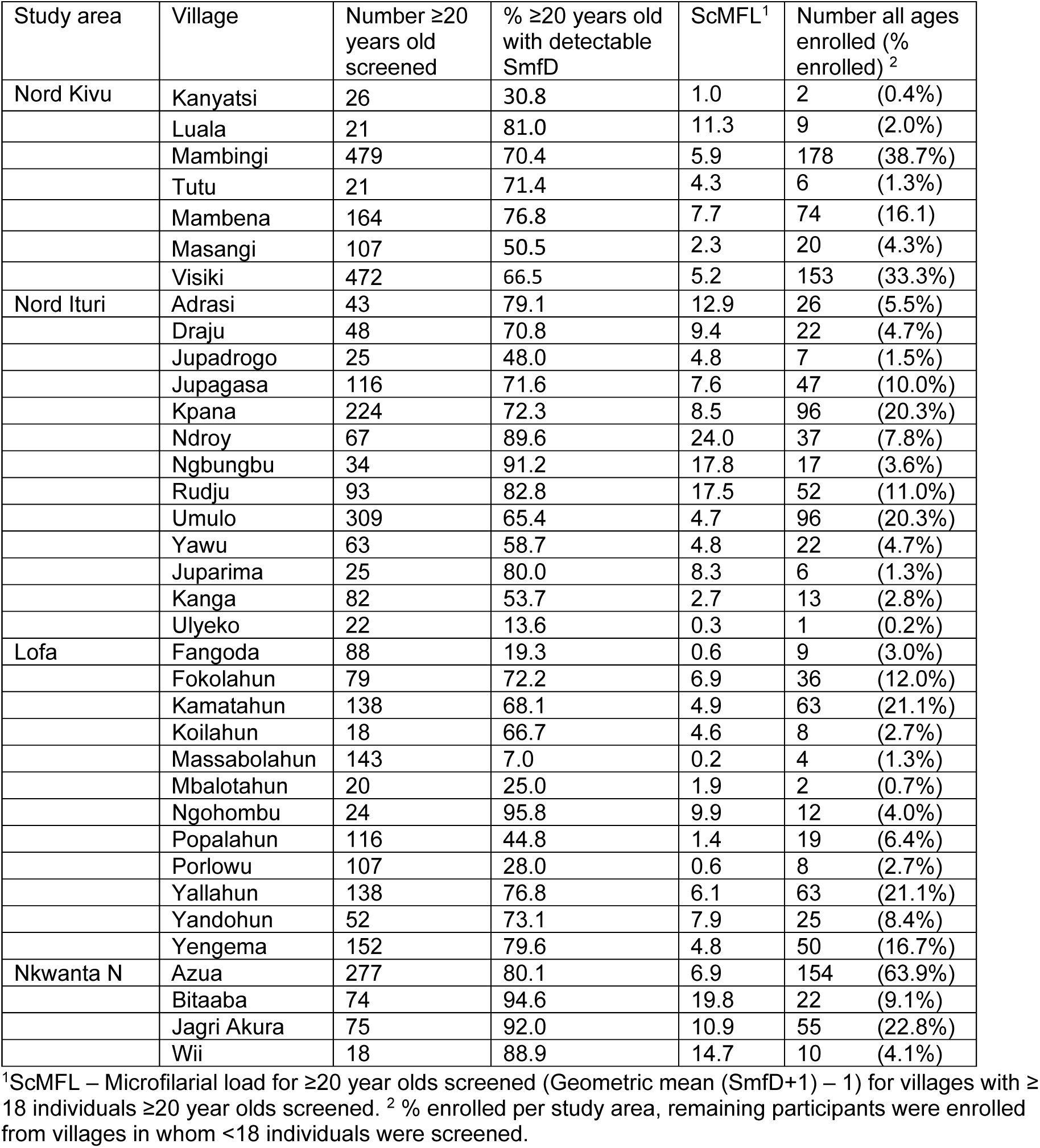
Community infection indicators among screened individuals ≥20 years by study area and village.

### Demographic and pre-treatment *O. volvulus* infection characteristics of enrolled study participants by study area

Table 3 shows the demographic and pre-treatment characteristics of enrolled study participants by study area and treatment arm.

**Table 3:**
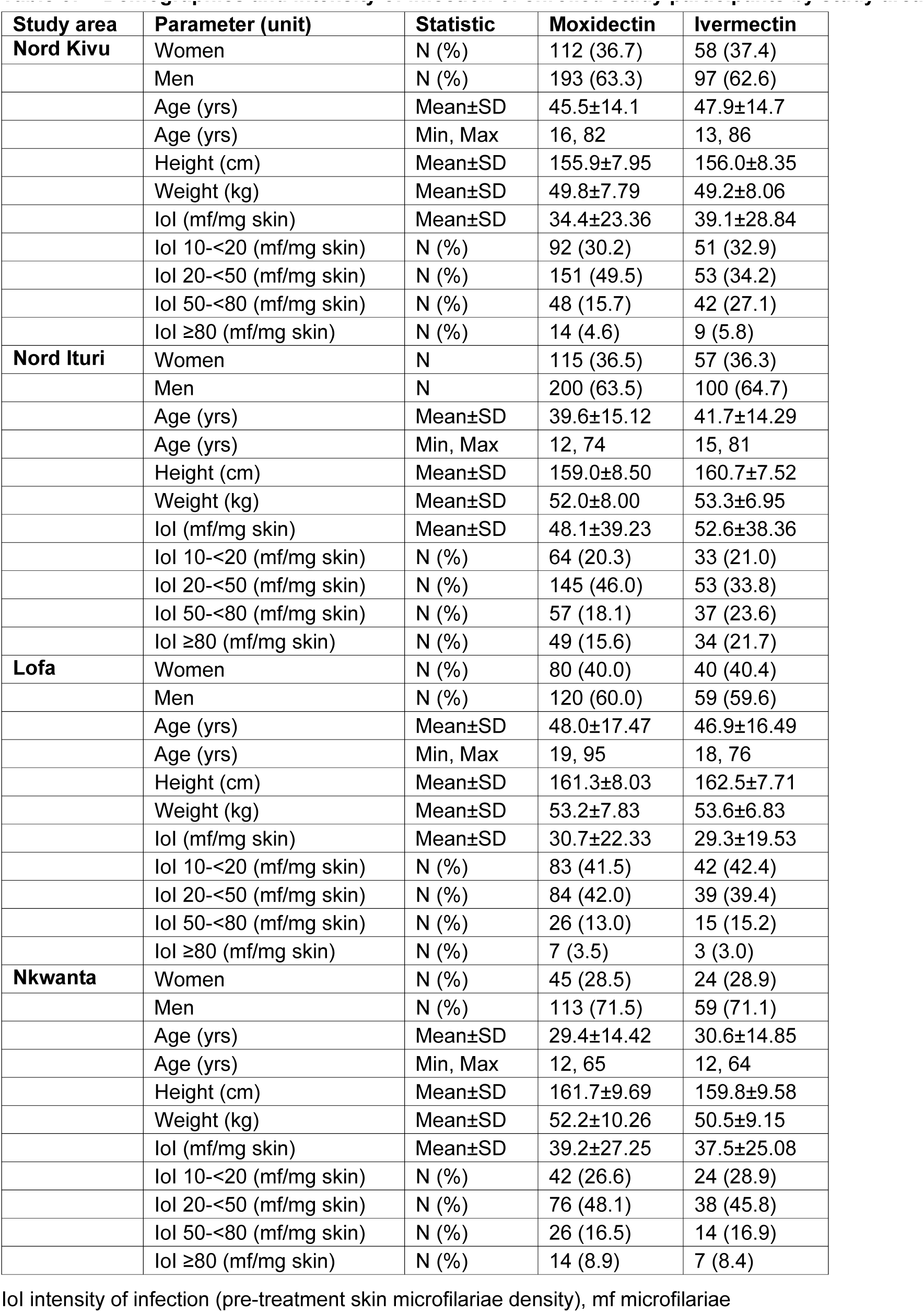
Demographics and intensity of infection of enrolled study participants by study area.

### Effect of moxidectin and ivermectin on skin microfilariae density

The number of individuals treated and with follow up data as well as descriptive statistics of SmfD by IoI category across all study areas and by study area are available in S1 Table S2 to Table S6. Across the study, follow-up rates in the two treatment arms were >99% at month 1, 98-99% at month 6, 96-97% at month 12 and, due to the protocol amendment, 78% at month 18 [30].

#### Post treatment skin microfilariae densities by study area

Fig 4 shows the SmfD before and 1, 6, 12 and 18 months post treatment by study area. The percentage reduction in geometric mean SmfD at baseline to month 1 ranged from 99.7%-100% in the moxidectin and from 97.4%-97.9% in the ivermectin treatment arm. Table 4 shows the moxidectin – ivermectin treatment difference at each post-treatment evaluation by study area.

**Fig 4:**
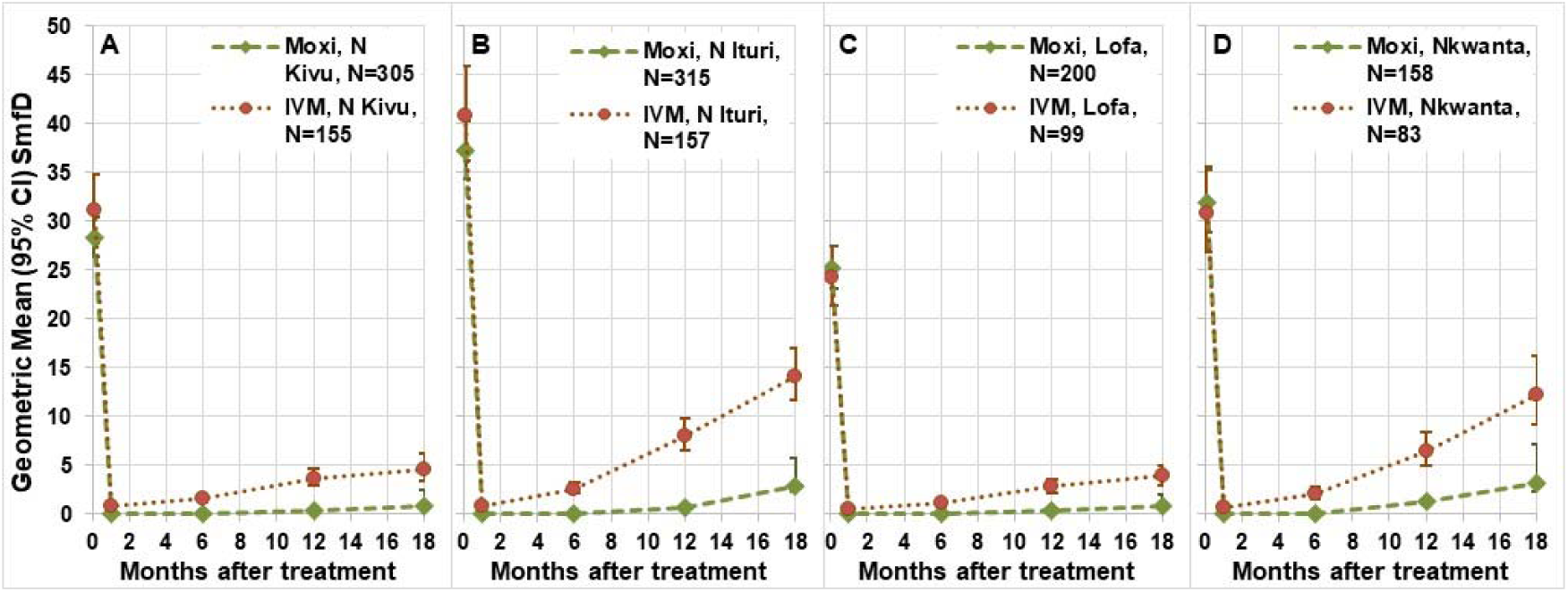
Geometric mean (95% CI) SmfD before and after treatment by study area. N Kivu Nord Kivu, N Ituri Nord Ituri, Lofa Lofa County, Nkwanta Nkwanta North district

**Table 4:** Treatment difference 1, 6, 12 and 18 months post-treatment by study area.

| Time post treatment | DRC Nord Kivu | DRC Nord Ituri | Liberia Lofa | Ghana Nkwanta |
| --- | --- | --- | --- | --- |
| Month 1 | 98.6 % | 83.7 % | 97.1 % | 110.7 % |
| Month 6 | 100.4 % | 98.8 % | 98.8 % | 103.2 % |
| Month 12 | 92.9 % | 90.1 % | 86.8 % | 84.5 % |
| Month 18 | 84.1 % | 78.0 % | 78.2 % | 78.2 % |

SmfD increased from month 1 onward in the ivermectin arm and from month 6 onward in the moxidectin treatment arm. These increases were higher in Nord Ituri and Nkwanta North than in Nord Kivu and Lofa County. The similar mean IoI for Nord Kivu and Nkwanta North and similar percentage of participants with ≥50 mf/mg skin IoI suggest that the differences in SmfD increases between the study areas is unlikely to be due solely to differences in IoI among participants (Fig 4). Pairwise study area comparison of the SmfD 12 months after treatment with adjustment for IoI showed that the efficacy of IVM was similar in Nord Kivu and Lofa County as well as in Nord Ituri and Nkwanta district whereas the efficacy in Nord Kivu and Lofa County was significantly higher than that in Nord Ituri and the Nkwanta district (p≤0.0002). The efficacy of moxidectin was similar in Nord Kivu, Nord Ituri and Lofa County and significantly higher in these three areas than in Nkwanta district (p<0.006).

Treatment difference: difference between adjusted geometric means of IVM and Moxi in percentage of the adjusted geometric means in the ivermectin treatment arm obtained from the linear model. Treatment differences >100% are due to low values of SmfD in the moxidectin arm that are log-transformed to calculate geometric mean differences. For summaries of raw SmfD, geometric means, adjusted means and geometric means and adjusted difference of means and geometric means see S1 Table S2 to Table S6.

#### Post treatment skin microfilariae densities by pre-treatment intensity of infection

The difference in SmfD post-treatment between the moxidectin and ivermectin treatment arm increased with increasing IoI category (Fig 5). Twelve months post-treatment, (i.e. the time of the next treatment during an annual moxidectin treatment strategy) the treatment difference across study areas was 74.1%, 84.2%, 90.0% and 95.4% among those with IoI of 10-<20, ≥20-<50, ≥50-<80, ≥80, respectively (p<0.0001). In the linear model, both the interaction of treatment and time and the interaction of treatment and IoI category confirmed (p<0.0001) that the difference in SmfD post-treatment between the moxidectin and ivermectin treatment arms increased over time as well as between IoI categories.

**Fig 5:**
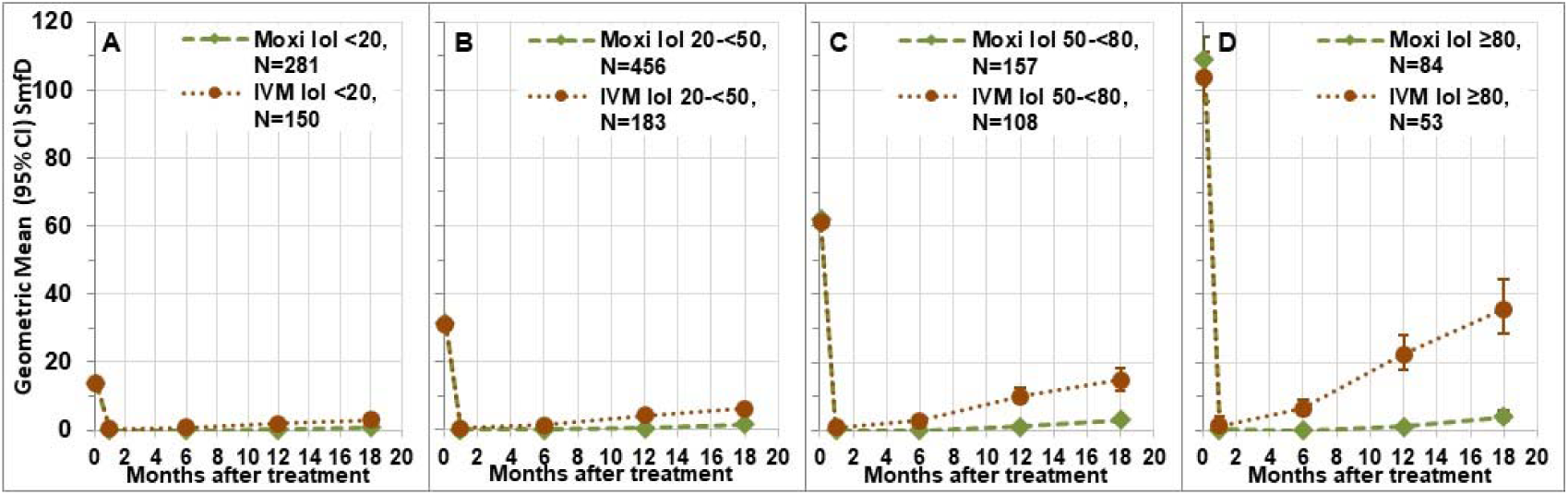
Geometric mean (95% CI) SmfD by intensity of infection category across study areas. SmfD: skin microfilariae density, moxi: moxidectin, IVM: ivermectin, N: number treated (for number with follow up data at each time point, see S1 Table S2).

The analysis by study area showed the same trend towards higher treatment differences among participants in the higher IoI categories as the across study area analysis. In all study areas, participants in the highest IoI category benefitted more from moxidectin treatment than those in the lowest IoI category (Fig 6, Table 5). Descriptive statistics for the SmfD by IoI category across and by study area are provided in S1 Table S2 to Table S6.

**Fig 6:**
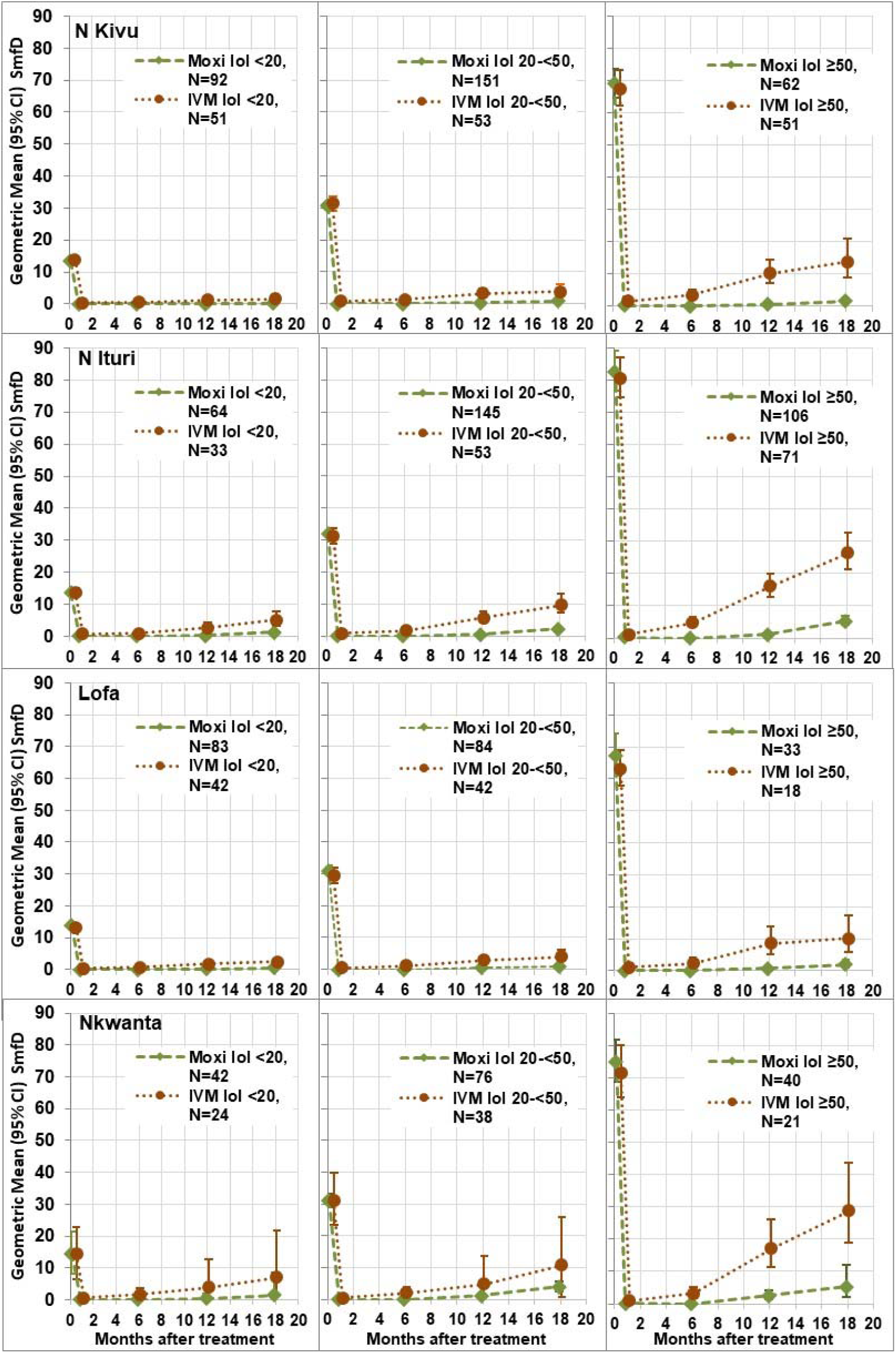
Geometric mean (95% CI) SmfD by intensity of infection category and study area. Moxi: moxidectin, IVM: ivermectin, N: number treated (for number with follow up data at each time point, see S1 Tables A2-A5), IoI intensity of infection: <20: 10-<20 mf/mg skin; 20-50: 20-<50 mf/mg skin, ≥50: ≥50 mf/mg skin pre-treatment.

**Table 5:**
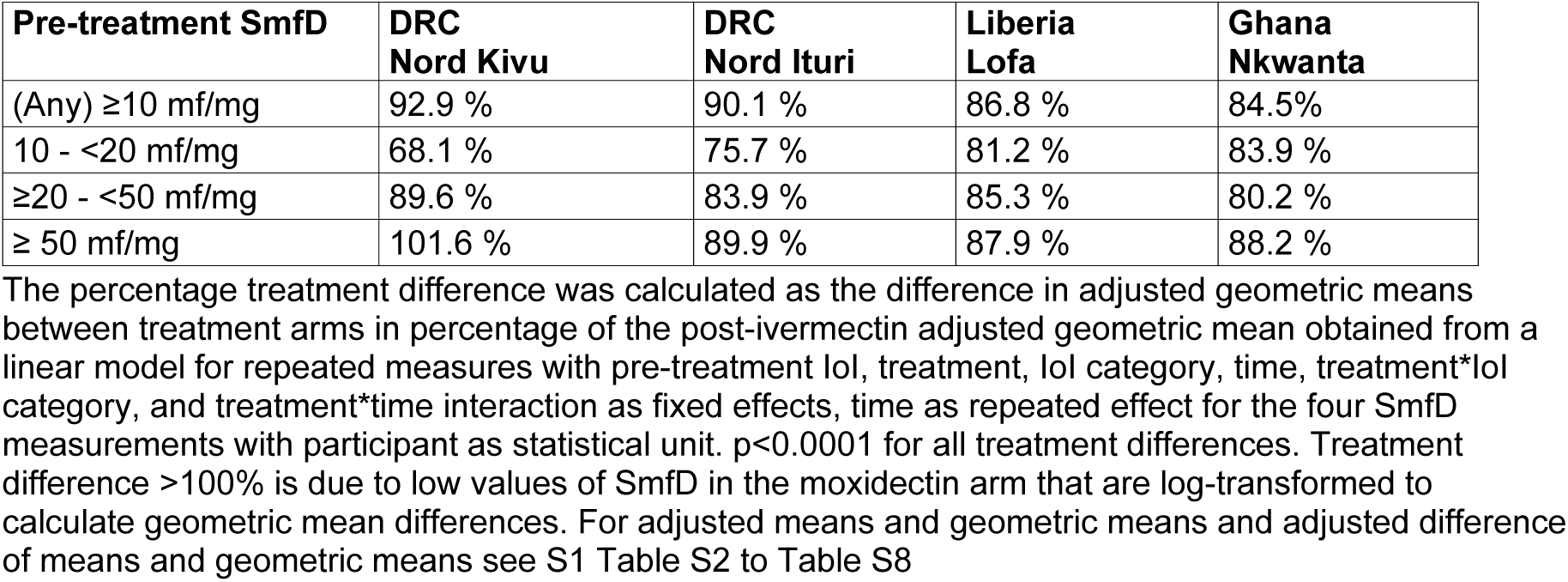
Treatment difference 12 months post-treatment by study area across and by intensity of infection.

#### Percentage of participants with undetectable SmfD post treatment by study area and pre-treatment intensity of infection

Across all study areas, 151/959 (15.7%) of participants with data at both 1 and 6 months after moxidectin treatment had lower SmfD 6 months (0.02±0.11 mf/mg, range 0-1.23 mf/mg) than 1 month (0.53±1.03, range 0.05-8.37 mf/mg) after treatment resulting in 136 participants achieving undetectable SmfD only at 6 months. Since a high percentage of moxidectin treated participants retained undetectable SmfD from month 1 to month 6, this resulted in a higher percentage of participants with undetectable SmfD from Month 1 to Month 6 (Fig 7) than at month 1 (84%, [30]). Such an increase was not observed in the ivermectin treatment arm, but 107/490 (21.8%) of participants in the ivermectin treatment arm with data at both 1 and 6 months had lower SmfD 6 months (4.50±8.66 mf/mg, range 0-43.1 mf/mg) than 1 month (7.81±13.36 mf/mg, range 0.08-53.6 mf/mg) after treatment.

**Fig 7:**
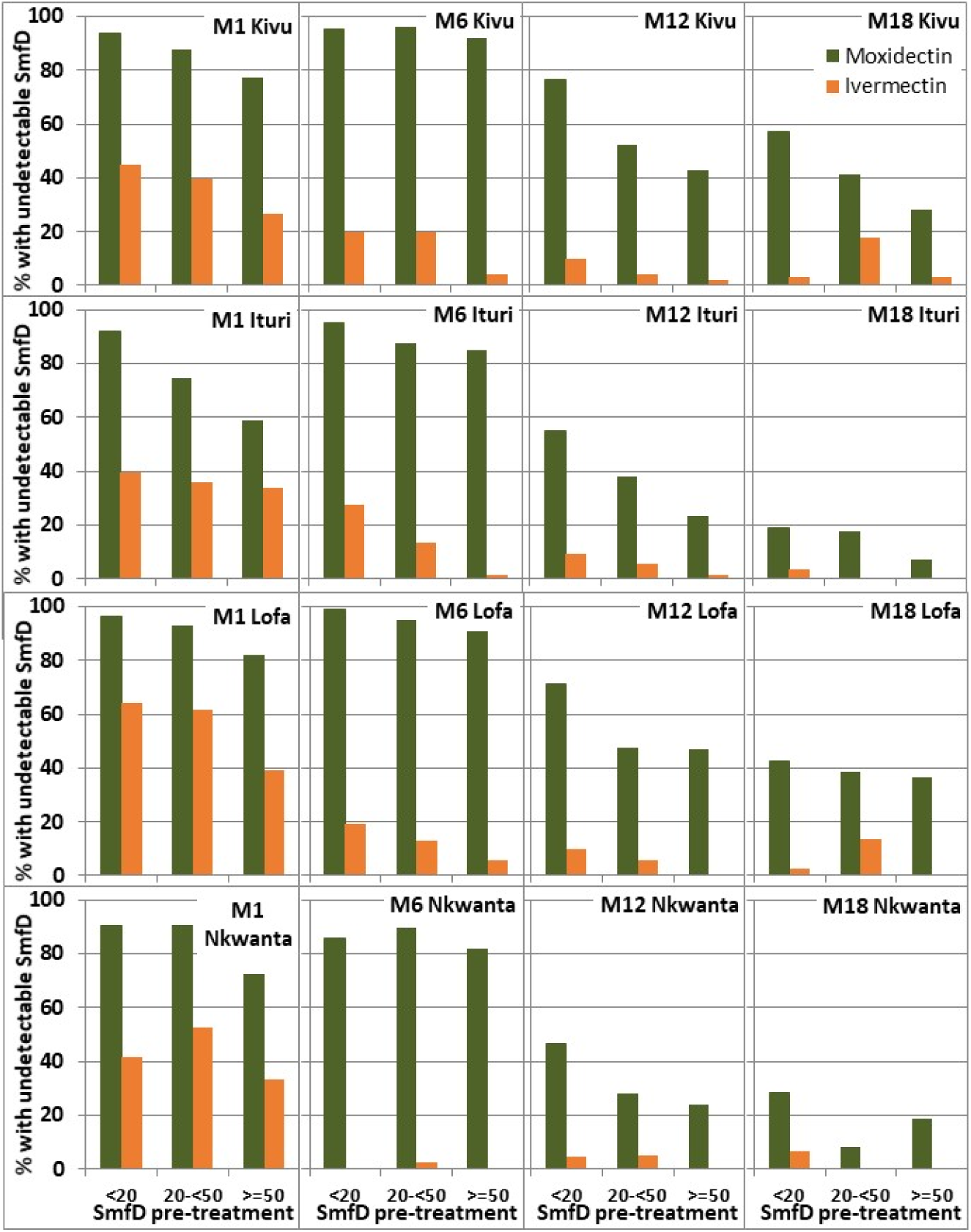
Percentage of participants with undetectable SmfD by study area, pre-treatment SmfD and post-treatment time. M1, M6, M12, M18: month 1, 6, 12 18 after treatment (for numbers treated see Fig 6, for numbers with follow up data at each time point, see S1 Table S2 to Table S6)

It is noteworthy that small changes in SmfD from one follow up time point to the other may not reflect actual changes in SmfD but rather the accuracy of SmfD detection. This has a relatively low impact on analyses based on mean SmfD, but can have a significant impact on categorical analyses such as the percentage with undetectable SmfD. The majority of moxidectin treated participants with detectable SmfD at Month 1 and undetectable SmfD at Month 6 had <0.25 mf/mg skin at Month 1 (Fig 8).

**Fig 8:**
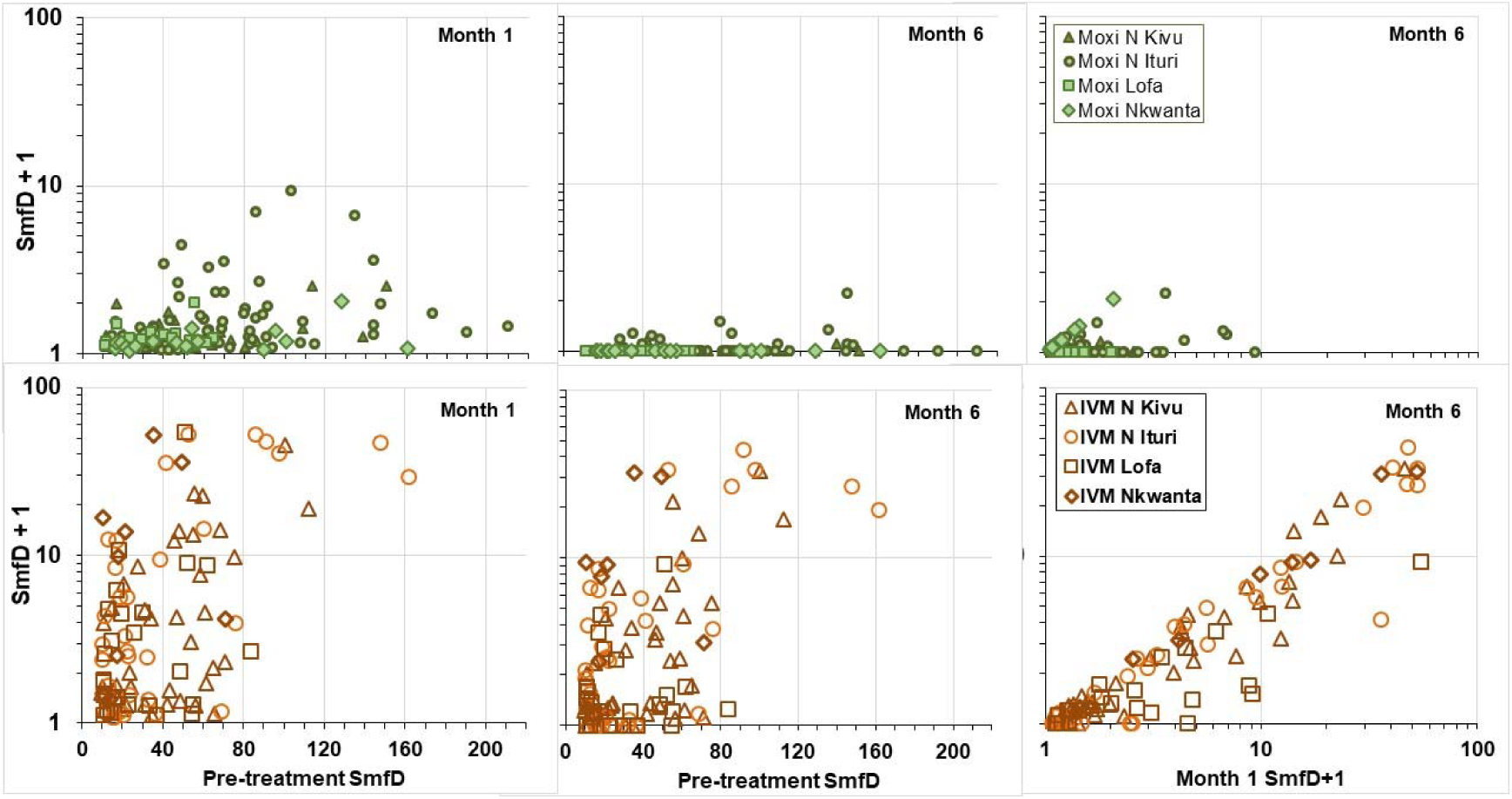
Skin microfilariae levels pre-treatment, at month 1 and at month 6 among participants who had detectable levels 1 month after treatment. SmfD: Skin microfilariae density, to allow displaying these data on a logarithmic scale, 1 was added to the SmfD values at Month 1 and 6.

#### Duration of undetectable levels of SmfD

The number of individuals in each study area with and without undetectable SmfD (UD) and those not evaluable for this analysis because SmfD data were missing for at least one of the relevant time points is provided in S1 Table S9. The logistic model derived odds and their lower and upper limits are provided in S1 Table S10 and displayed in S1 Fig S1 and the odds ratios are provided in S1 Table S11. Table S9 shows that there were not zero participants with undetectable SmfD in the ivermectin arm in the higher IoI categories which resulted in the logistic model not providing odds and odds ratios (Table S10 and Table S11).

Fig 9 shows the logistic model derived odds of study participants across all study areas to have UD from 1-6, 1-12 and 1-18 months after treatment with moxidectin or ivermectin by the four IoI categories defined for the across study area analysis.

**Fig 9:**
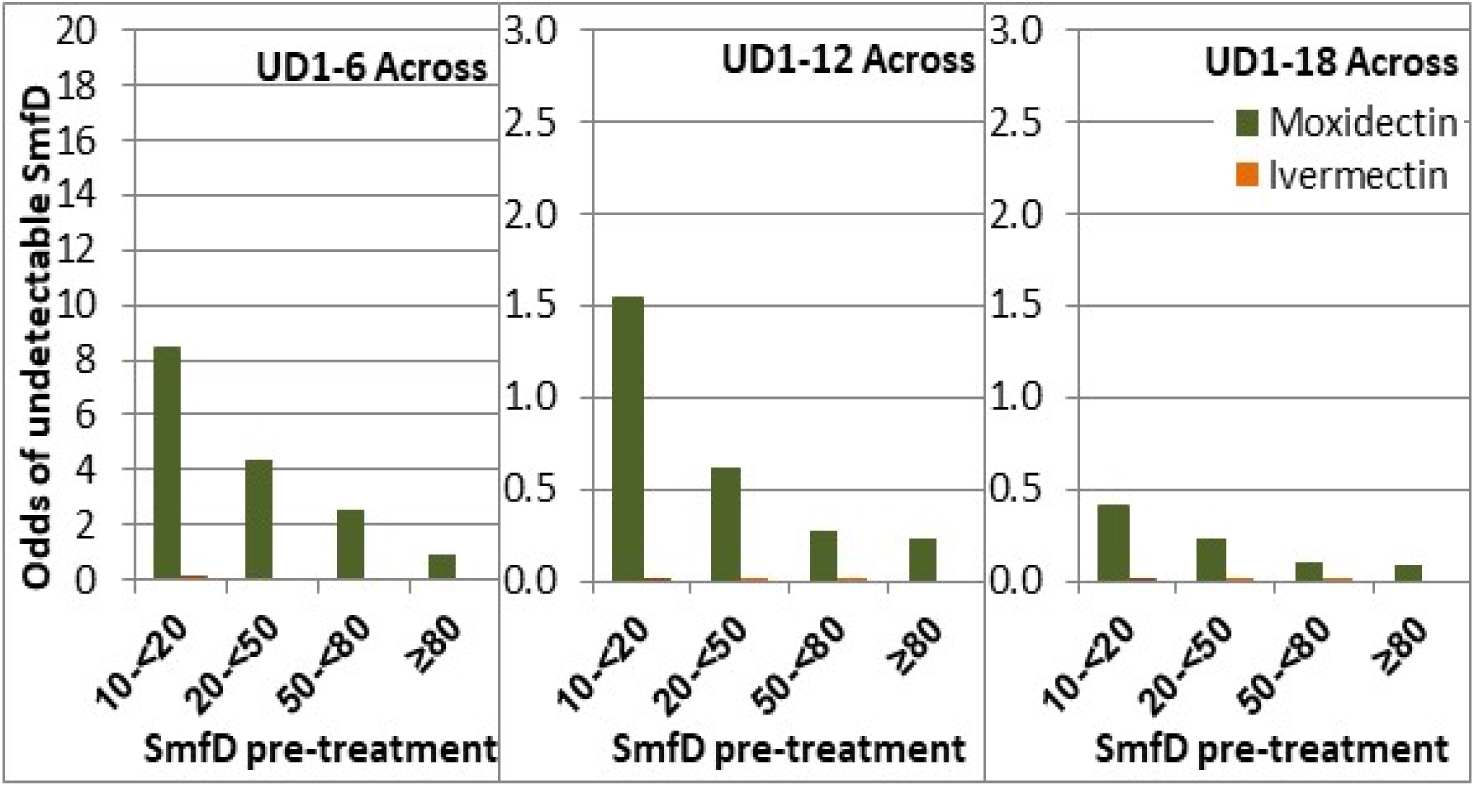
Odds for undetectable SmfD from 1 to 6, 12 or 18 months by IoI across study areas. SmfD: skin microfilariae density, UD1-6, UD 1-12, UD 1-18 undetectable levels of SmfD at month 1 and 6, at month 1, 6 and 12, and at month 1, 6, 12 and 18, respectively (for number with follow up data at each relevant time point, see Table A8).

The odds of moxidectin treated participants to have UD of any duration were significantly higher than those of ivermectin treated participants in each IoI category (p<0.0002 for each IoI category) but the difference in odds between the moxidectin and ivermectin arms was not significantly different between IoI categories (p>0.8 for the interaction between treatment and IoI which was discarded from the models). The highest odds in the ivermectin treatment arm were 0.13 for individuals with <20 mf/mg IoI to have UD 1-6.

The same trends were observed in the analysis by study area. Moxidectin treated participants had significantly higher odds of having UD1-6, 1-12 and 1-18 than ivermectin treated participants (p< 0.008 for each study area) where data was sufficient to allow the comparison in marginal means. The difference in odds for moxidectin and ivermectin treated participants was not significantly different between the different IoI categories (treatment – IoI interaction not significant).

### Inter-individual variability in response to moxidectin and ivermectin

#### ‘Suboptimal microfilariae response’ to treatment

Awadzi et al. considered ≥60% reduction of SmfD from pre-treatment to day 8 as indicating an ‘adequate parasite response’. In the first study comparing the effect of moxidectin and ivermectin on SmfD, 3/45 ivermectin treated participants had an SmfD reduction on day 7 or 8 after treatment lower than that and were referred to as ‘suboptimal microfilariae responders’ (SOMR) [29]. In this study, the first post-treatment SmfD measurement occurred 1 month after treatment. SmfD decreases from Day 8 to 1-2 months after a single dose of ivermectin [29,39,40]. Therefore, the criterion used here to assess SOMR was reduction of ≤ 80% in SmfD from pre-treatment to month 1 (SOMR80).

In the moxidectin treatment arm, no individual met the SOMR80 criteria: the % reduction in SmfD from pre-treatment to month 1 ranged from 91.9 -100% in Nord Kivu, 94.2 - 100% in Nord Ituri, 96.9% - 100% in Lofa County, 99.0 - 100% in the Nkwanta District. All 42 individuals with <99% SmfD reduction at month 1 had >99% SmfD reduction at month 6.

In the ivermectin arm, the percentage reduction in SmfD from pre-treatment to month 1 ranged from 51.0% – 100% in Nord Kivu, 2% – 100% in Nord Ituri, -4.7% – 100% in Lofa County and - 49.6% – 100% in Nkwanta District. Across study areas, 8.9% of ivermectin treated met the SOMR80 criteria. Fig 10 shows the SmfD at each post treatment time point for the 44 individuals meeting the SOMR80 criteria. Among these, 21 also met the ‘suboptimal response’ (SOR) criteria (see below), including 4/14 in Nord Kivu, 10/18 in Nord Ituri, 1/5 in Lofa Country and 6/7 in Nkwanta District. The data do not suggest a dependency of the percentage of SOMR individuals on IoI or study area. This is also not the case when stricter criteria for SOMR are used, i.e. reduction of ≤90% in SmfD from pre-treatment to month 1 (SOMR90) (Table 6).

**Fig 10:**
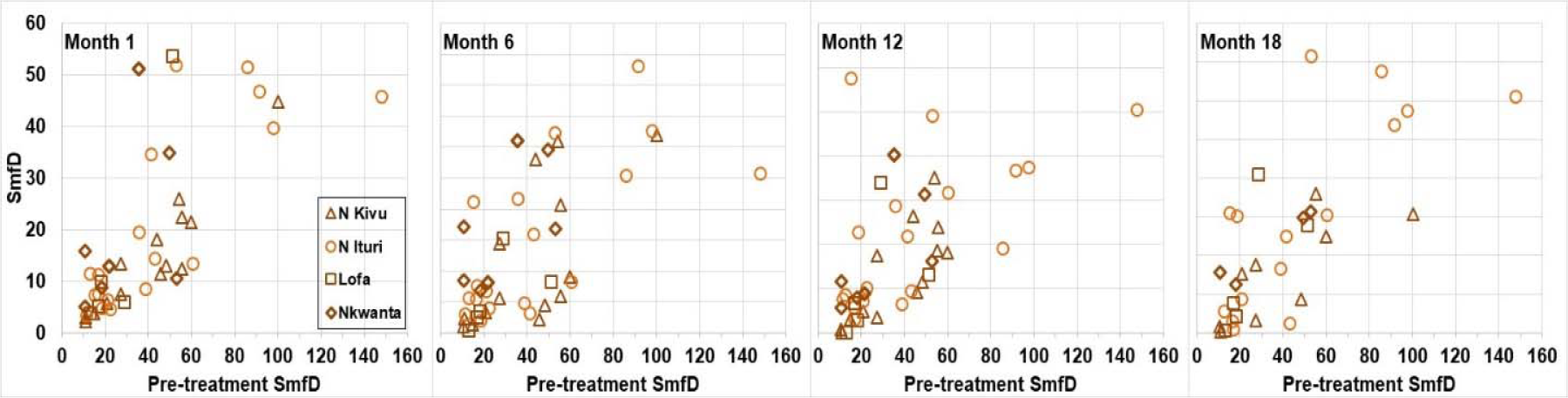
SmfD 1, 6, 12 and 18 month post-treatment in participants with ‘suboptimal microfilariae response’ to ivermectin as indicated by ≤80% reduction of skin microfilariae levels from pre-treatment to 1 month after treatment’.

**Table 6:** Percentage of participants with ‘suboptimal microfilariae response’.

| lol (mf/mg) | DRC Nord Kivu<br>SOMR80 (SOMR90) |  | DRC Nord Ituri<br>SOMR80 (SOMR90) |  | Liberia Lofa County<br>SOMR80 (SOMR90) |  | Ghana Nkwanta district<br>SOMR80 (SOMR90) |  |
| --- | --- | --- | --- | --- | --- | --- | --- | --- |
|  | Moxi | IVM | Moxi | IVM | Moxi | IVM | Moxi | IVM |
| (Any) $\geq 10$ | 0 (0) | 9.2 (15.6) | 0 (0) | 11.5 (14.7) | 0 (0) | 5.1 (14.1) | 0 (0) | 8.4 (13.3) |
| 10 - <20 | 0 (0) | 5.9 (14.0) | 0 (0) | 18.2 (24.2) | 0 (0) | 7.1 (16.7) | 0 (0) | 12.5 (16.7) |
| $\geq 20$ - <50 | 0 (0) | 11.3 (15.4) | 0 (0) | 11.3 (13.2) | 0 (0) | 2.6 (10.3) | 0 (0) | 5.3 (13.2) |
| $\geq 50$ | 0 (0) | 10.2 (18.4) | 0 (0) | 8.5 (11.3) | 0 (0) | 5.6 (16.7) | 0 (0) | 9.5 (9.5) |

SOMR80: suboptimal microfilariae response based on $\leq 80\%$ SmfD reduction from pre-treatment to month 1 post treatment. SOMR90 suboptimal microfilariae response based on $\leq 90\%$ SmfD reduction from pre-treatment to month 1 post treatment

#### Minimum SmfD in participants with detectable SmfD at all post-treatment measurements to Month 12

Among the 965 moxidectin and 491 ivermectin treated participants with Month 1 as well as Month 6 and/or 12 SmfD measurement, 19 (1.9%) and 257 (52.3%) of participants had detectable SmfD at all evaluated time points (range after moxidectin treatment 0.1 – 1.2 mf/mg, range after ivermectin treatment 0.1 – 36.8 mf/mg), respectively. The % of participants who did not achieve undetectable SmfD after treatment was highest among participants in the IoI ≥50 mf/mg category (Table 8). There was no correlation between pre-treatment SmfD and the minimum detectable post-treatment SmfD among either moxidectin or ivermectin treated individuals (Fig 11).

**Fig 11:**
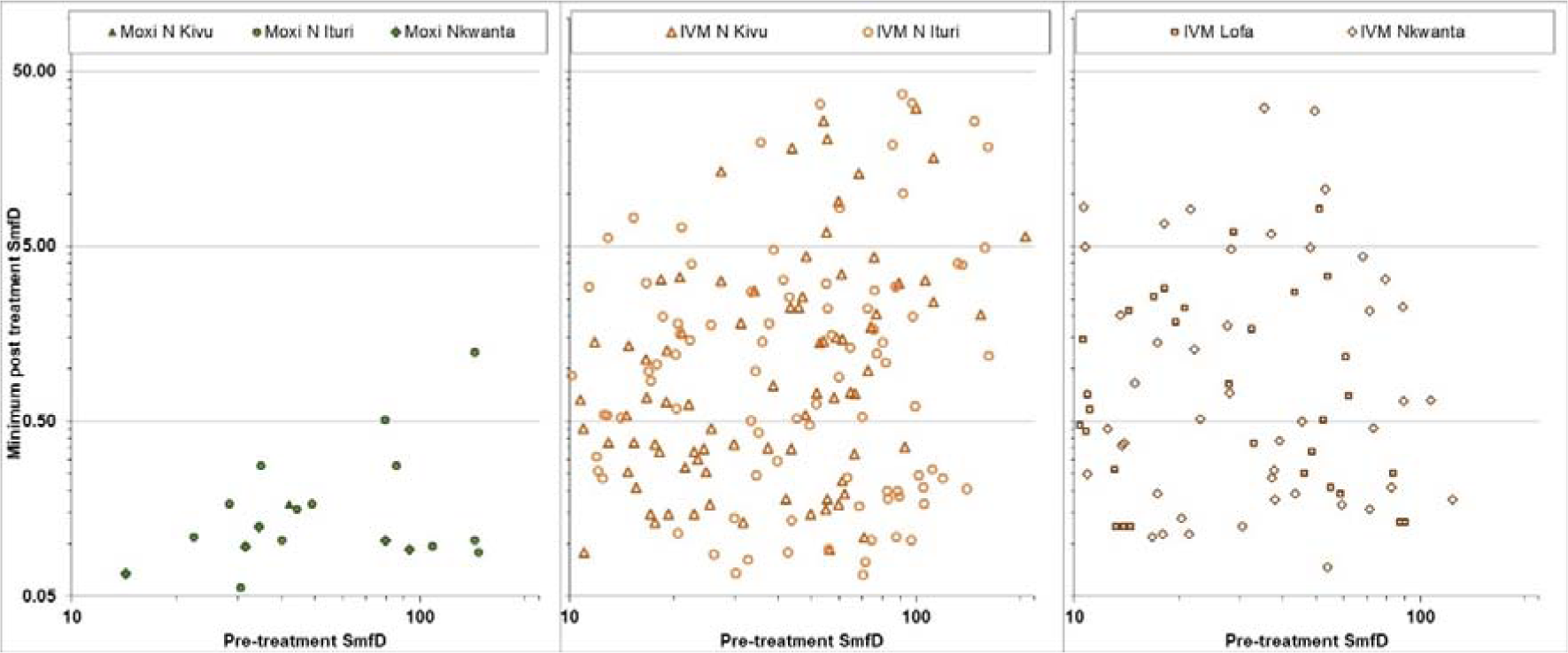
Minimum SmfD for participants with detectable SmfD at all post-treatment evaluations. Left: moxidectin treated participants, center: ivermectin treated participants from DRC, right: ivermectin treated participants from Lofa country and Nkwanta North

#### ’Suboptimal response’ to treatment

A number of criteria have been developed for ’suboptimal response’ (SOR) to ivermectin, i.e. individuals in whom the initial decrease in SmfD is followed by an increase ‘larger than expected’ or considered ‘typical’ (for overview of criteria and references see supplementary information in [30]). Using the SOR criterion of SmfD at month 12 post treatment of >40% of the pre-treatment SmfD proposed during the first investigation into SOR [39, 41], the percentage of participants with SOR to moxidectin (10/947, 1.1%) was substantially lower than the percentage of participants with SOR to ivermectin (88/480, 18.3%). Among those with SOR to ivermectin, 21 also met the SOMR80 criteria, including 4/14 in Nord Kivu, 10/18 in Nord Ituri, 1/5 in Lofa Country and 6/7 in Nkwanta District. For both drugs, the percentage of SOR differed between study areas but did not increase with IoI category (Table 9). The independence of SOR of IoI is further supported by the across study area analysis of individuals with IoI ≥80: Among the 83 moxidectin treated and 52 ivermecin treated individuals with IoI ≥80 and month 12 data, 0.0% and 21.2%, respectively, met the SOR criterion. Fig 12 shows for each participant meeting the SOR criterion the SmfD at each measurement by pre-treatment SmfD.

**Fig 12:**
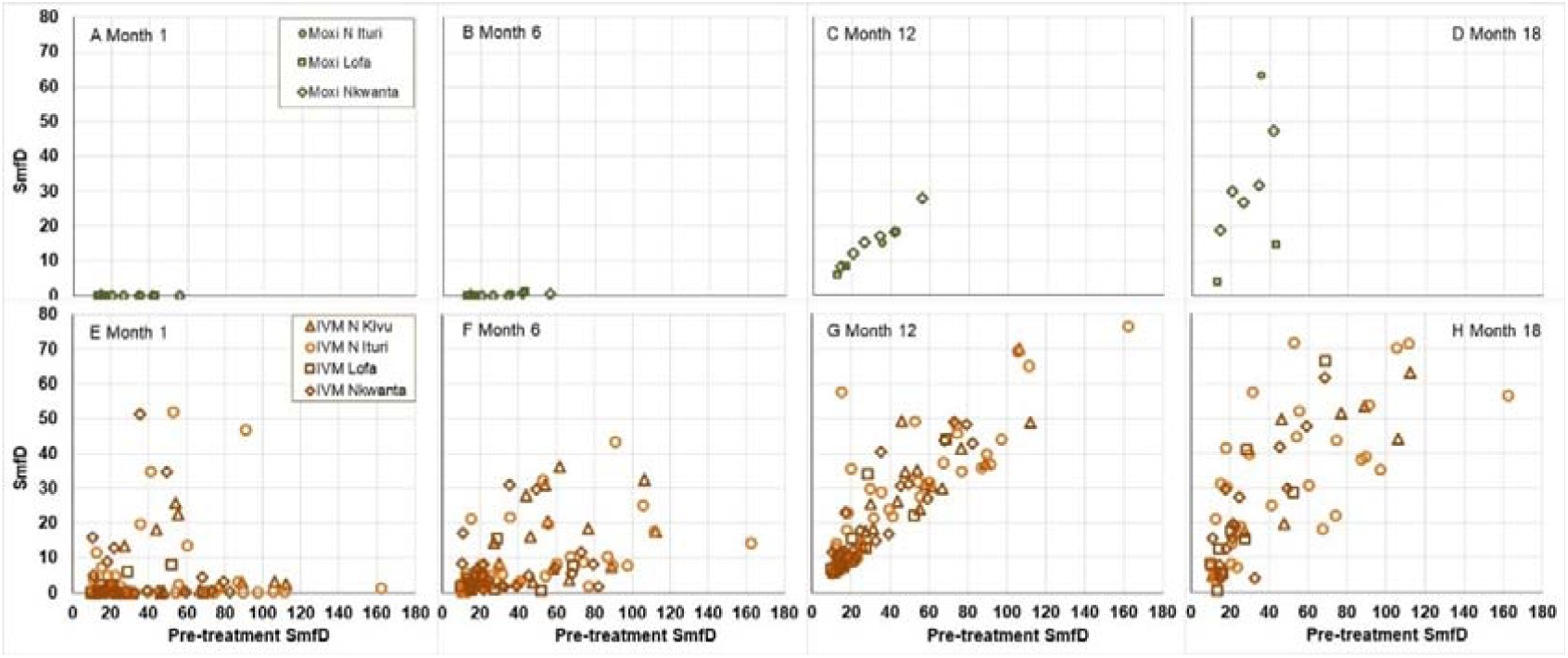
SmfD at Month 1, 6, 12 and 18 vs pre-treatment SmfD for the 10 moxidectin and 88 ivermectin treated participants with ‘suboptimal response’ participants with ‘suboptimal response’.

## Discussion and Conclusions

Moxidectin treatment induced a maximum SmfD reduction from pre-treatment by 100% in 98.1% of the 978 treated participants and by 99.1%-99.9% in the remaining 19 participants. In conjunction with the minimum post-treatment SmfD of these 19 individuals (0.1-1.2 mf/mg), the data did not suggest any dependency of SmfD reduction on IoI (Fig 11) or study area. Among the 494 ivermectin treated individuals, the by-study area analysis identified 5.1% - 11.5%

SOMR80 (individuals with a SmfD 1 month after treatment reduced by ≤80% from pre-treatment SmfD). Between 31.3% and 58.0% of ivermectin treated individuals with Month 1 and Month 6 and/or Month 12 measurements had detectable SmfD at all post-treatment time points. The data did not suggest a dependency of the extent of initial SmfD reduction on IoI or study area and the SmfD in most of these individuals were too high to be attributed to the sensitivity of the skin snip method [42] (Table 7, Fig 11).

**Table 7:** Skin microfilariae density 1 month after ivermectin among participants without and with ‘suboptimal microfilariae response’.

| lol (mf/mg) | DRC Nord Kivu |  | DRC Nord Ituri |  | Liberia Lofa County |  | Ghana Nkwanta district |  |
| --- | --- | --- | --- | --- | --- | --- | --- | --- |
|  | OMR | SOMR80 | OMR | SOMR80 | OMR | SOMR80 | OMR | SOMR80 |
| | AM±SD | AM±SD | AM±SD | AM±SD | AM±SD | AM±SD | AM±SD | AM±SD |
| (Any) $\geq 10$ | 0.9±2.2 | 14.7±11.5 | 0.8±2.6 | 21.3±18.1 | 0.6±1.4 | 15.7±21.3 | 0.6±1.3 | 19.9±16.9 |
| 10 - <20 | 0.4±0.6 | 3.0±0.8 | 0.3±0.5 | 7.6±3.3 | 0.3±0.6 | 6.2±3.1 | 0.3±0.5 | 9.9±5.5 |
| $\geq 20$ - <50 | 0.6±1.0 | 11.5±4.4 | 0.5±0.7 | 14.7±11.2 | 0.5±1.0 | 6.0 | 0.7±1.6 | 32.1±27.2 |
| $\geq 50$ | 1.9±3.6 | 25.4±11.9 | 1.3±3.8 | 41.5±14.5 | 1.3±2.6 | 53.6 | 0.8±1.4 | 22.7±17.1 |

OMR: Optimal microfilariae response to ivermectin based on $>80\%$ reduction of SmfD from pre-treatment by Month 1, SOMR80 suboptimal microfilariae response to ivermectin based on $\leq 80\%$ reduction of SmfD from pretreatment by Month 1, AM arithmetic mean SmfD at Month 1, SD standard deviation of arithmetic mean SmfD at Month 1

**Table 8:** Participants with detectable SmfD and SmfD data at 1 and 6 and/or 12 months post- treatment.

|  | DRC Nord Kivu |  | DRC Nord Ituri |  | Liberia Lofa County |  | Ghana Nkwanta district |  |
| --- | --- | --- | --- | --- | --- | --- | --- | --- |
| Iol | Moxi | IVM | Moxi | IVM | Moxi | IVM | Moxi | IVM |
| (mf/mg) | n (%) | n (%) | n (%) | n (%) | n (%) | n (%) | n (%) | n (%) |
| (Any) $\geq 10$ | 1 (0.3) | 88 (57.9) | 13 (4.2) | 91 (58) | 0 | 33 (33.3) | 5 (3.2) | 45 (54.2) |
| 10 - <20 | 0 | 24 (27.4) | 0 | 16 (48.5) | 0 | 12 (28.6) | 1 (2.4) | 13 (54.2) |
| $\geq 20$ - <50 | 1 (0.7) | 29 (56.9) | 7 (4.9) | 29 (54.7) | 0 | 11 (28.2) | 2 (2.7) | 18 (47.4) |
| $\geq 50$ | 0 | 35 (70.0) | 6 (5.7) | 46 (64.8) | 0 | 10 (55.6) | 2 (5.1) | 14 (66.7) |

SmfD Skin microfilariae density

**Table 9:** Percentage of participants with ‘suboptimal response’ to moxidectin or ivermectin.

|  | DRC Nord Kivu |  | DRC Nord Ituri |  | Liberia Lofa County |  | Ghana Nkwanta district |  |
| --- | --- | --- | --- | --- | --- | --- | --- | --- |
| Iol | Moxi | IVM | Moxi | IVM | Moxi | IVM | Moxi | IVM |
| (mf/mg) | % | % | % | % | % | % | % | % |
| (Any) $\geq 10$ | 0 | 12.1 | 0.3 | 23.7 | 1.6 | 10.8 | 3.9 | 28.0 |
| 10 - <20 | 0 | 3.9 | 0 | 33.3 | 2.6 | 12.2 | 2.4 | 34.8 |
| $\geq 20$ - <50 | 0 | 14.0 | 0.7 | 20.8 | 1.3 | 8.3 | 5.3 | 23.7 |
| $\geq 50$ | 0 | 18.8 | 0 | 21.4 | 0.0 | 12.5 | 2.6 | 28.6 |

Despite the percentages of ivermectin treated individuals who were SOMR80s or did not have undetectable SmfD at any post-treatment time, the reduction from pre-treatment to Month 1 in the ivermectin treatment group ranged from 97.4-97.9%. This is similar to the 98-99% reduction at 1-2 months post treatment derived from meta-analysis of the aggregate data from 26 clinical and community single ivermectin dose studies conducted in ivermectin-naïve individuals in Liberia, Mali, Ghana, Cameroon, Senegal, Togo, Ethiopia, Sierra Leone, Cote d’Ivoire and Guatemala [40] and the ≈98% reduction 15 days after treatment among ivermectin-naïve and ivermectin multi-treated individuals from the Nkam and Mbam valley in Cameroon [43]. Given that the skin microfilariae are the reservoir for parasite transmission, including inter-individual heterogeneity in SmfD reduction may be important when modelling the long term effect of CDTI on time to parasite elimination [44, 45].

In contrast to the initial SmfD reduction, the repopulation of the skin with microfilariae depended on IoI, consistent with the fact that the IoI reflects the number of reproductively active macrofilariae in an individual, our previous analysis which considered only the two IoI categories used to according to which individuals had been stratified for randomization (10-20 mf/mg, ≥20) [30] and analyses of data obtained after the first ivermectin treatment of individuals from Cameroon [46]. The dependency was modest after moxidectin but pronounced after ivermectin treatment (Fig 5, Fig 6). This difference resulted in individuals with higher IoI benefitting relatively more from moxidectin treatment than individuals with lower IoI as indicated by across-study treatment differences 12 months post treatment of 74.1%, 84.2%, 90.0% and 95.4% among those with IoI of 10-<20, ≥20-<50, ≥50-<80, ≥80.

The by-study area analysis showed that repopulation of the skin with microfilariae, evident at month 6 in the ivermectin but only at month 12 in the moxidectin group, was more extensive among individuals in Nord Ituri and Nkwanta North than individuals in Nord Kivu and Lofa Country (Fig 4). This difference was not driven exclusively by differences in IoI distribution among the participants in the different study areas (Fig 6). The percentages of SOR to ivermectin were higher in Nord Ituri and Nkwanta North District (23.7% and 28%, resp.) than in Nord Kivu and Lofa County (12.1% and 10.8%, resp.). The percentage of SOR was not consistently higher among individuals with IoI ≥50 than among those with IoI 10-20 and thus not driven by the IoI.

The first study into differences in skin repopulation rates between individuals and communities was initiated in 2001 to evaluate potential causes of persistent significant microfilaridermia in some individuals in communities in the Lower Black Volta and Pru river basins after >10 years of ivermectin MDA. A clinical study excluded pharmacokinetic reasons and concluded that individuals with SmfD 3 and 12 months after treatment exceeding 6% and 40% of pre- ivermectin treatment SmfD (calculated per 4 mg skin based on four snips), respectively, and embryonic stages in the uteri of macrofilariae 90 days after treatment harbor parasites who respond as expected to ivermectin’s ‘microfilaricidal effect’ but are non-responsive or sub-optimally responsive (SOR) to ivermectin’s ‘embryostatic effect’ [39, 41]. Ivermectin’s ‘embryostatic effect’ results in skin repopulation with microfilariae starting weeks after ivermectin has been cleared [47]. Ivermectin has a half life of <1 day [48]. Investigation in Cameroon of skin repopulation rates in ivermectin-naïve individuals from the Nkam valley where CDTI had not yet been initiated and multi-treated individuals from the Mbam Valley with ongoing CDTI showed that skin repopulation rates were higher in multi-treated than ivermectin-naïve individuals to 80 days but comparable to 180 days after ivermectin treatment [43]. The investigators of further studies in the Lower Black Volta, Pru, and Daka river basins suggested that detectable levels of skin microfilariae (based on two snips) 90 days after treatment, SmfD (calculated per snip) 12 months after treatment exceeding 100% of pre-treatment SmfD and/or presence of embryonic stages of microfilariae in the uteri of macrofilariae 90 days after treatment may indicate emerging parasite resistance to ivermectin’s embryostatic effect [49, 50].

Provided that microfilariae produced by SOR *O. volvulus* have the same fitness as microfilariae from *O. volvulus* responding ‘normally / as expected’ to the embryostatic effect of ivermectin (i.e. the same probability of being ingested by the vector, developing into infective larvae, being transmitted and developing into reproductively capable macrofilariae if transmitted), earlier skin repopulation with the microfilariae from SOR than ‘normally responding’ parasites could result in increasing prevalence of SOR parasites if their appearance in the skin occurs while vectors are abundant. This emphasizes the importance of timing and frequency of CDTI relative to transmission seasons. Optimization of CDTI timing was one of the recommendations emerging from the APOC consultations on ‘Strategic Options and Alternative Treatment Strategies for Accelerating Onchocerciasis Elimination in Africa [21]. Such considerations would be less important if moxidectin rather than ivermectin was used for onchocerciasis control and elimination.

Our data on prevalence of SOR to ivermectin also highlight the need for longitudinal analysis of available data to determine whether or not SOR prevalence is increasing with CDTI duration.

Should this be the case, tools suitable for large scale monitoring of SOR prevalence would be needed to guide national program decisions. Development of tools based on genetic correlates of SOR is one approach [51], but the need for phenotypic characterization via serial skin snipping or embryogramms remains a laborious and costly challenge [52].

CDTI had not yet been implemented in the areas where the study was conducted. Consequently, ivermectin selection pressure may not be the explanation for the differences in skin repopulation between the study areas that cannot be attributed to differences in IoI distribution. It takes 10-15 months for a transmitted larvae to mature into a microfilariae- releasing macrofilaria [1]. Therefore, it is unlikely that differences in endemicity and new infections between study areas are responsible for the differences in skin microfilariae repopulation to month 12. Consideration thus needs to be given to the possibility that ‘ivermectin-naïve’ *O. volvulus* populations in different areas differ in their susceptibility to ivermectin (and to a much lower degree to moxidectin). To date, this possibility has not yet been considered in analyses of data from different geographic areas [40, 53]. The hypothesis that parasite populations in different geographic areas have different susceptibility to ivermectin and different inter-individual variability of this susceptibility is compatible with *O. volvulus* biology.

Bottlenecks within the life cycle of *O. volvulus* severely restrict the fraction of the progeny of one generation which contribute to the next generation and genetic drift could result in different genetic make up of different parasite populations. Genome-wide association analyses of the *O. volvulus* nuclear genome of phenotypically characterized macrofilariae from Ghana [49, 50] and Cameroon [43,54,55] suggest significant genetic differentiation between the parasite populations investigated [51].

While our results are consistent with the hypothesis of different ivermectin susceptibility of different ivermectin-naïve parasite populations, they cannot be considered as supporting this hypothesis without further investigation: while the areas where we recruited participants were CDTI-naïve, areas not far away (e.g. for Nkwanta district other villages within the Oti river basin, in the Pru River basins and across the border in Togo) had been benefitting from CDTI for many years. The parasites in our study participants could belong to the same parasite population as the parasites in individuals in nearby areas which have been undergoing CDTI for many years. The population genetic structure of parasites obtained in the SOR studies in Ghana [49, 50] suggests that parasites in individuals in the Pru, Daka and Black Volta/Tombe river basins belong to the same transmission zone in which parasites interbreed, enabled through vector or people movement ([52]). Without genetic analysis of the parasites from the Kpasa subdistrict in Nkwanta North it cannot be excluded that they belong to that same interbreeding parasite population. If that is the case and if CDTI selects for SOR, this selection in the Pru, Daka and Black Volta/Tombe river basins could be reflected in the parasite population in our study participants. Long range transmission of parasites through infected/infective vector movement was a challenge for the Onchocerciasis Control Programme in West Africa, resulting in extensions of the original programme area [56–59]. Recently long range vector movement has been implicated in renewed transmission of *O. volvulus* in the Comoé valley of Burkina Faso, although movement of infected people could not be excluded [60, 61].

The differences in efficacy of ivermectin and moxidectin in terms of the initial decrease in SmfD and in terms of maintaining undetectable or low SmfD through 18 months post treatment result in higher odds of moxidectin compared to ivermectin treated participants having undetectable SmfD at month 1-6, month 1-12 and month 1-18 in each study area and each IoI category.

Given that the skin microfilariae are the reservoir of transmission, this confirms our conclusion from the across study area analysis that moxidectin treatment may be particularly advantageous for achieving elimination of *O. volvulus* transmission in areas where infection prevalence remains high despite long-term CDTI, where transmission seasons are long or have two peaks, where elimination programmes face operational barriers to implementing at least annual CDTI and where SOR to ivermectin has been observed [30]. This conclusion is supported by the results of modelling the effect of annual or biannual community directed treatment with moxidectin compared to annual or biannual CDTI [62].

## Data Availability

Participants consented to publication of summaries of the results, not to sharing of their individual data. Consequently, the Sponsor (WHO) and the authors do not have the participants permission to make individual participant data publicly available. Individuals wanting to analyze the data should contact the Sponsor (via the corresponding author or tdr at who.int) or Medicines Development for Global Health to which WHO has licensed the data (via mark.sullivan at medicinesdevelopment.com). Requests should include the objectives, data analysis plan and plans to obtain applicable Ethics Committee approvals and involve the investigators (co-authors on this manuscript) and commitment to not share the data with anybody else.

## Acknowledgements

We acknowledge the contribution to data collection in Liberia of Mr. Mawolo Kpawor, who died in October 2016. We acknowledge Dr Fatorma Bolay who directed study center creation and preparation, community engagement, parasitologist training and management of the Liberian study site (see [30]). He died in March 2021. For others whose contribution to the study we want to acknowledge, see [30].

We are particularly grateful to all study participants for their co-operation.

The authors alone are responsible for the views expressed which do not necessarily represent the views, decisions or policies of the institutions with which the authors are affiliated.

## Data availability

Participants consented to publication of summaries of the results, not to sharing of their individual data. Consequently, the Sponsor (WHO) and the authors do not have the participants’ permission to make individual participant data publicly available. Individuals wanting to analyze the data should contact the Sponsor (via the corresponding author) or Medicines Development for Global Health to which WHO has licensed the data (via). Requests should include the objectives, data analysis plan and plans to obtain applicable Ethics Committee approvals and involve the investigators (co-authors on this manuscript) and commitment to not share the data with anybody else.

In view of the restriction to underlying data sharing, the figures and tables provided in the manuscript have been complemented with detailed output of the statistical analyses in the Supporting Information S1.

## Financial Disclosure

WHO/TDR funded this study, utilizing contributions from the WHO African Programme for Onchocerciasis Control (APOC), 6.3 million $US from Wyeth and following its acquisition by Pfizer, Pfizer, and WHO/TDR donor countries. Wyeth provided drug for this study and contributed to the study protocol. Wyeth prepared the submissions to the Ministries of Health and provided data management services until July 3, 2011. Pfizer was not further involved in this study in any way, including data verification or analysis and has not commented on this manuscript.

## Competing interests

I have read the journal’s policy and the authors of this manuscript have the following competing interests: ACK and CMH are staff of WHO which funded the work of all co-authors on the study whose data are analysed here through its department UNICEF/UNDP/World Bank/WHO Special Programme for Research and Training in Tropical Diseases (TDR).

## Copyright

© 2020 World Health Organization. Licensee Public Library of Science.

This is an open access article distributed under the Creative Commons Attribution IGO License, which permits unrestricted use, distribution, and reproduction in any medium, provided the original work is properly cited. https://creativecommons.org/licenses/by/3.0/igo/. In any use of this article, there should be no suggestion that WHO endorses any specific organization, products or services. The use of the WHO logo is not permitted. This notice should be preserved along with the article’s original URL.

## Supporting information S1

**Table S1.** GPS coordinates of participant villages, research center and towns in the vicinity.

| Study area | Village where participants lived | Research Center | Towns for reference | Latitude | Longitude |
| --- | --- | --- | --- | --- | --- |
| Nord Kivu, DRC | Mambena |  |  | 0.418306 | 29.225528 |
| Nord Kivu, DRC | Mambingi |  |  | 0.36656 | 29.27531 |
| Nord Kivu, DRC | Visiki |  |  | 0.37709 | 29.23835 |
| Nord Kivu, DRC |  | Butembo |  | 0.11405 | 29.30180 |
| Nord Kivu, DRC |  |  | Beni | 0.49901 | 29.45275 |
| Nord Ituri, DRC | Adrasi |  |  | 2.123217 | 30.969 |
| Nord Ituri, DRC | Draju |  |  | 2.118517 | 30.966067 |
| Nord Ituri, DRC | Jupadrogo |  |  | 2.1278 | 30.966067 |
| Nord Ituri, DRC | Jupagasa |  |  | 2.121 | 30.982133 |
| Nord Ituri, DRC | Juparima |  |  | 2.0868 | 30.955917 |
| Nord Ituri, DRC | Kanga |  |  | 2.074233 | 30.954283 |
| Nord Ituri, DRC | Kpana |  |  | 2.1114 | 30.96585 |
| Nord Ituri, DRC | Ndroy |  |  | 2.103917 | 30.974417 |
| Nord Ituri, DRC | Ngbungbu |  |  | 2.120117 | 30.97925 |
| Nord Ituri, DRC | Rudju |  |  | 2.124517 | 30.9835 |
| Nord Ituri, DRC | Ulyeko |  |  | 2.13545 | 30.956383 |
| Nord Ituri, DRC | Umulo |  |  | 2.131033 | 30.972117 |
| Nord Ituri, DRC | Yawu |  |  | 2.125017 | 30.965883 |
| Nord Ituri, DRC |  | Rethy |  | 2.07500 | 30.92165 |
| Nord Ituri, DRC |  |  | Bunia | 1.57428 | 30.23973 |
| Nord Ituri, DRC |  |  | Logo | 2.183167 | 30.927333 |
| Nord Ituri, DRC |  |  | Mahagi | 2.30547 | 30.97616 |
| Lofa County, Lib. | Kamatahun |  |  | 8.1368 | -10.297133 |
| Lofa County, Lib. | Yallahun |  |  | 8.131033 | -10.3124 |
| Lofa County, Lib. | Yandohun |  |  | 8.105333 | -10.35805 |
| Lofa County, Lib. | Yengma |  |  | 8.170283 | -10.307933 |
| Lofa County, Lib. | Fokolahun |  |  | 8.15935 | -10.274517 |
| Lofa County, Lib. |  | Bolahun |  | 8.23092 | -10.16064 |
| Lofa County, Lib. |  |  | Foya | 8.39492 | -10.21516 |
| Lofa County, Lib. |  |  | Lofa | 8.19112 | -9.72327 |
| Lofa County, Liberia |  |  | Voinjama | 8.42020 | -9.75393 |
| Nkwanta Dist., Ghana | Azua |  |  | 8.57208 | 0.40642 |
| Nkwanta Dist., Ghana | Bitaaaba |  |  | 8.51159 | 0.31829 |
| Nkwanta Dist., Ghana | Jagri Akura |  |  | 8.53148 | 0.35278 |
| Nkwanta Dist., Ghana | Wii |  |  | 8.51974 | 0.33389 |
| Ghana |  | Hohoe |  | 7.15185 | 0.473829 |
| Nkwanta District, Ghana |  |  | Kpasa | 8.48962 | 0.30428 |
| Nkwanta District, Ghana |  |  | Nkwanta | 8.26031 | 0.52211 |
| Ghana |  |  | Wiae | 8.32275 | -0.16192 |
| Ghana |  |  | Asubende | 7.91667 | -1.16667 |
| Ghana |  |  | Kyingakrom | 8.10122 | -2.05871 |

**Table S2.** SmfD pre-treatment, 1, 6, 12 and 18 months post treatment across all study areas by intensity of infection pre-treatment.

| Pre-Tx Iol (mf/mg) | Treatment Group | N | Raw Mean (SD) | Raw 95% CI | Raw Median (1Q,3Q) | Raw Min., Max. | Geometric Mean (95%CI) | Overall p-value (a) |
| --- | --- | --- | --- | --- | --- | --- | --- | --- |
| **Pre-treatment SmfD** |  |  |  |  |  |  |  |  |
| 10 - <20 | Moxidectin | 281 | 13.9 (2.9) | [13.6,14.3] | 13.0 (11.0,16.0) | 9.0,19.0 | 13.7 [13.4,14.0] | <0.0001 |
|  | Ivermectin | 150 | 14.1 (2.9) | [13.6,14.5] | 14.0 (11.0,17.0) | 10.0,19.0 | 13.8 [13.3,14.2] |  |
| 20 - <50 | Moxidectin | 456 | 32.3 (8.6) | [31.5,33.1] | 31.0 (24.0,40.0) | 20.0,49.0 | 31.2 [30.4,32.0] |  |
|  | Ivermectin | 183 | 31.9 (8.3) | [30.7,33.1] | 31.0 (25.0,38.0) | 20.0,49.0 | 30.9 [29.7,32.1] |  |
| 50 - <80 | Moxidectin | 157 | 62.4 (7.9) | [61.2,63.7] | 62.0 (56.0,69.0) | 50.0,79.0 | 61.9 [60.7,63.2] |  |
|  | Ivermectin | 108 | 62.0 (8.6) | [60.3,63.6] | 60.5 (55.0,69.0) | 50.0,79.0 | 61.4 [59.8,63.0] |  |
| ≥80 | Moxidectin | 84 | 113.7 (37.0) | [105.6,121.7] | 100.5 (89.0,135.0) | 80.0,299.0 | 109.1 [102.8,115.8] |  |
|  | Ivermectin | 53 | 107.4 (32.7) | [98.4,116.5] | 93.0 (87.0,112.0) | 80.0,238.0 | 103.7 [96.7,111.3] |  |
| **Month 1** |  |  |  |  |  |  |  |  |
| 10 - <20 | Moxidectin | 281 | 0.0 (0.1) | [-0.0,0.0] | 0.0 (0.0,0.0) | 0.0,1.0 | 0.0 [-0.0,0.0] | <0.0001 |
|  | Ivermectin | 150 | 0.9 (2.4) | [0.6,1.3] | 0.0 (0.0,1.0) | 0.0,16.0 | 0.4 [0.3,0.6] |  |
| 20 - <50 | Moxidectin | 452 | 0.0 (0.2) | [0.0,0.0] | 0.0 (0.0,0.0) | 0.0,3.0 | 0.0 [0.0,0.0] |  |
|  | Ivermectin | 183 | 1.7 (5.4) | [0.9,2.5] | 0.0 (0.0,1.0) | 0.0,51.0 | 0.6 [0.4,0.8] |  |
| 50 - <80 | Moxidectin | 157 | 0.1 (0.3) | [0.0,0.1] | 0.0 (0.0,0.0) | 0.0,3.0 | 0.0 [0.0,0.1] |  |
|  | Ivermectin | 106 | 3.2 (8.9) | [1.5,5.0] | 0.0 (0.0,2.0) | 0.0,54.0 | 0.9 [0.6,1.3] |  |
| ≥80 | Moxidectin | 83 | 0.5 (1.3) | [0.2,0.8] | 0.0 (0.0,0.0) | 0.0,8.0 | 0.2 [0.1,0.4] |  |
|  | Ivermectin | 53 | 6.2 (13.8) | [2.4,10.0] | 0.0 (0.0,3.0) | 0.0,52.0 | 1.5 [0.8,2.5] |  |
| **Month 6** |  |  |  |  |  |  |  |  |
| 10 - <20 | Moxidectin | 275 | 0.0 (0.1) | [-0.0,0.0] | 0.0 (0.0,0.0) | 0.0,2.0 | 0.0 [-0.0,0.0] | <0.0001 |
|  | Ivermectin | 150 | 1.3 (2.6) | [0.9,1.8] | 1.0 (0.0,1.0) | 0.0,21.0 | 0.8 [0.6,1.0] |  |
| 20 - <50 | Moxidectin | 449 | 0.0 (0.1) | [0.0,0.0] | 0.0 (0.0,0.0) | 0.0,1.0 | 0.0 [0.0,0.0] |  |
|  | Ivermectin | 181 | 2.6 (4.4) | [2.0,3.3] | 1.0 (0.0,3.0) | 0.0,31.0 | 1.5 [1.2,1.8] |  |
| 50 - <80 | Moxidectin | 154 | 0.0 (0.1) | [-0.0,0.0] | 0.0 (0.0,0.0) | 0.0,1.0 | 0.0 [-0.0,0.0] |  |
|  | Ivermectin | 107 | 5.1 (7.2) | [3.8,6.5] | 2.0 (1.0,7.0) | 0.0,36.0 | 2.8 [2.1,3.5] |  |
| ≥80 | Moxidectin | 84 | 0.0 (0.2) | [0.0,0.1] | 0.0 (0.0,0.0) | 0.0,1.0 | 0.0 [0.0,0.1] |  |
|  | Ivermectin | 53 | 10.5 (10.2) | [7.7,13.3] | 7.0 (3.0,17.0) | 0.0,43.0 | 6.5 [4.7,8.9] |  |
| **Month 12** |  |  |  |  |  |  |  |  |
| 10 - <20 | Moxidectin | 270 | 0.4 (1.0) | [0.2,0.5] | 0.0 (0.0,0.0) | 0.0,8.0 | 0.2 [0.1,0.3] | <0.0001 |
|  | Ivermectin | 148 | 3.4 (6.0) | [2.5,4.4] | 2.0 (1.0,4.0) | 0.0,58.0 | 1.9 [1.6,2.4] |  |
| 20 - <50 | Moxidectin | 440 | 1.1 (2.7) | [0.9,1.4] | 0.0 (0.0,1.0) | 0.0,18.0 | 0.5 [0.4,0.6] |  |
|  | Ivermectin | 177 | 7.0 (8.3) | [5.7,8.2] | 4.0 (2.0,10.0) | 0.0,49.0 | 4.2 [3.5,4.9] |  |
| 50 - <80 | Moxidectin | 154 | 2.3 (4.4) | [1.6,3.0] | 0.0 (0.0,2.0) | 0.0,28.0 | 1.0 [0.7,1.3] |  |
|  | Ivermectin | 103 | 15.2 (13.0) | [12.6,17.7] | 11.0 (6.0,22.0) | 0.0,49.0 | 10.0 [8.0,12.3] |  |
| ≥80 | Moxidectin | 83 | 2.6 (4.5) | [1.6,3.5] | 1.0 (0.0,2.0) | 0.0,20.0 | 1.2 [0.8,1.6] |  |
|  | Ivermectin | 52 | 28.6 (18.1) | [23.5,33.6] | 24.5 (15.0,38.5) | 1.0,77.0 | 22.5 [17.9,28.1] |  |
| **Month 18** |  |  |  |  |  |  |  |  |
| 10 - <20 | Moxidectin | 270 | 0.4 (1.0) | [0.2,0.5] | 0.0 (0.0,0.0) | 0.0,8.0 | 0.2 [0.1,0.3] | <0.0001 |
|  | Ivermectin | 148 | 3.4 (6.0) | [2.5,4.4] | 2.0 (1.0,4.0) | 0.0,58.0 | 1.9 [1.6,2.4] |  |
| 20 - <50 | Moxidectin | 440 | 1.1 (2.7) | [0.9,1.4] | 0.0 (0.0,1.0) | 0.0,18.0 | 0.5 [0.4,0.6] |  |
|  | Ivermectin | 177 | 7.0 (8.3) | [5.7,8.2] | 4.0 (2.0,10.0) | 0.0,49.0 | 4.2 [3.5,4.9] |  |
| 50 - <80 | Moxidectin | 154 | 2.3 (4.4) | [1.6,3.0] | 0.0 (0.0,2.0) | 0.0,28.0 | 1.0 [0.7,1.3] |  |
|  | Ivermectin | 103 | 15.2 (13.0) | [12.6,17.7] | 11.0 (6.0,22.0) | 0.0,49.0 | 10.0 [8.0,12.3] |  |
| ≥80 | Moxidectin | 83 | 2.6 (4.5) | [1.6,3.5] | 1.0 (0.0,2.0) | 0.0,20.0 | 1.2 [0.8,1.6] |  |
|  | Ivermectin | 52 | 28.6 (18.1) | [23.5,33.6] | 24.5 (15.0,38.5) | 1.0,77.0 | 22.5 [17.9,28.1] |  |

**Table S3.** SmfD pre-treatment, 1, 6, 12 and 18 months post treatment in Nord-Kivu (DRC) by intensity of infection pre-treatment.

| Pre-Tx Iol (mf/mg) | Treatment Group | N | Raw Mean (SD) | Raw 95% CI | Raw Median (1Q,3Q) | Raw Min., Max. | Geometric Mean [95%CI] | Overall p-value (a) |
| --- | --- | --- | --- | --- | --- | --- | --- | --- |
| Pre-treatment SmfD |  |  |  |  |  |  |  |  |
| 10 - <20 | Moxidectin | 92 | 13.7 (3.0) | [13.0,14.3] | 13.2 (11.0,16.2) | 9.7,19.5 | 13.4 [12.8,14.0] | <0.0001 |
|  | Ivermectin | 51 | 14.2 (3.3) | [13.3,15.2] | 14.7 (10.9,17.1) | 9.7,19.3 | 13.9 [13.0,14.8] |  |
| 20 - <50 | Moxidectin | 151 | 31.8 (8.7) | [30.4,33.2] | 31.3 (23.8,39.7) | 19.5,49.5 | 30.7 [29.3,32.1] |  |
|  | Ivermectin | 53 | 32.4 (8.3) | [30.1,34.6] | 30.8 (25.4,37.5) | 20.7,48.5 | 31.4 [29.2,33.6] |  |
| ≥50 | Moxidectin | 62 | 71.5 (21.5) | [66.0,76.9] | 65.4 (56.4,75.4) | 51.2,150.0 | 69.0 [64.7,73.6] |  |
|  | Ivermectin | 51 | 70.8 (27.9) | [63.0,78.7] | 61.3 (55.5,74.1) | 49.6,207.2 | 67.4 [62.1,73.2] |  |
| Month 1 |  |  |  |  |  |  |  |  |
| 10 - <20 | Moxidectin | 92 | 0.0 (0.1) | [-0.0,0.0] | 0.0 (0.0,0.0) | 0.0,1.0 | 0.0 [-0.0,0.0] | <0.0001 |
|  | Ivermectin | 51 | 0.5 (0.9) | [0.3,0.8] | 0.2 (0.0,0.6) | 0.0,3.8 | 0.4 [0.2,0.6] |  |
| 20 - <50 | Moxidectin | 148 | 0.0 (0.1) | [0.0,0.1] | 0.0 (0.0,0.0) | 0.0,0.8 | 0.0 [0.0,0.0] |  |
|  | Ivermectin | 53 | 1.8 (3.9) | [0.7,2.9] | 0.3 (0.0,0.9) | 0.0,18.2 | 0.8 [0.4,1.2] |  |
| ≥50 | Moxidectin | 62 | 0.1 (0.3) | [0.0,0.2] | 0.0 (0.0,0.0) | 0.0,1.5 | 0.1 [0.0,0.1] |  |
|  | Ivermectin | 49 | 4.3 (8.7) | [1.8,6.8] | 0.7 (0.0,3.1) | 0.0,44.8 | 1.6 [0.9,2.5] |  |
| Month 6 |  |  |  |  |  |  |  |  |
| 10 - <20 | Moxidectin | 89 | 0.0 (0.2) | [-0.0,0.1] | 0.0 (0.0,0.0) | 0.0,1.7 | 0.0 [-0.0,0.0] | <0.0001 |
|  | Ivermectin | 51 | 0.9 (1.3) | [0.5,1.2] | 0.4 (0.1,1.2) | 0.0,7.4 | 0.6 [0.4,0.9] |  |
| 20 - <50 | Moxidectin | 148 | 0.0 (0.0) | [0.0,0.0] | 0.0 (0.0,0.0) | 0.0,0.5 | 0.0 [0.0,0.0] |  |
|  | Ivermectin | 51 | 2.6 (4.9) | [1.3,4.0] | 0.9 (0.3,2.9) | 0.0,28.0 | 1.4 [0.9,2.0] |  |
| ≥50 | Moxidectin | 61 | 0.0 (0.1) | [-0.0,0.1] | 0.0 (0.0,0.0) | 0.0,0.7 | 0.0 [0.0,0.1] |  |
|  | Ivermectin | 50 | 6.8 (9.3) | [4.2,9.5] | 2.9 (0.9,8.6) | 0.0,36.3 | 3.5 [2.4,5.0] |  |
| Month 12 |  |  |  |  |  |  |  |  |
| 10 - <20 | Moxidectin | 90 | 0.1 (0.2) | [0.0,0.1] | 0.0 (0.0,0.0) | 0.0,1.3 | 0.1 [0.0,0.1] | <0.0001 |
|  | Ivermectin | 51 | 1.6 (1.7) | [1.1,2.1] | 0.9 (0.4,2.3) | 0.0,7.1 | 1.2 [0.8,1.6] |  |
| 20 - <50 | Moxidectin | 147 | 0.4 (0.8) | [0.3,0.6] | 0.0 (0.0,0.5) | 0.0,4.9 | 0.3 [0.2,0.4] |  |
|  | Ivermectin | 50 | 6.3 (9.6) | [3.6,9.0] | 2.7 (1.2,5.7) | 0.0,49.3 | 3.3 [2.2,4.6] |  |
| ≥50 | Moxidectin | 61 | 1.1 (2.4) | [0.5,1.7] | 0.2 (0.0,0.8) | 0.0,12.1 | 0.6 [0.3,0.8] |  |
|  | Ivermectin | 48 | 16.9 (15.5) | [12.4,21.4] | 14.7 (3.4,23.3) | 0.0,70.3 | 10.2 [7.1,14.4] |  |
| Month 18 |  |  |  |  |  |  |  |  |
| 10 - <20 | Moxidectin | 61 | 0.5 (1.4) | [0.2,0.9] | 0.0 (0.0,0.2) | 0.0,9.0 | 0.3 [0.1,0.4] | <0.0001 |
|  | Ivermectin | 34 | 2.3 (2.6) | [1.4,3.2] | 1.6 (0.4,2.7) | 0.0,12.1 | 1.6 [1.1,2.3] |  |
| 20 - <50 | Moxidectin | 87 | 1.4 (2.2) | [0.9,1.9] | 0.2 (0.0,2.3) | 0.0,10.7 | 0.8 [0.6,1.1] |  |
|  | Ivermectin | 34 | 7.9 (10.2) | [4.3,11.5] | 3.5 (0.4,15.2) | 0.0,49.8 | 3.8 [2.2,6.2] |  |
| ≥50 | Moxidectin | 53 | 3.4 (6.0) | [1.8,5.1] | 1.2 (0.0,3.3) | 0.0,26.5 | 1.6 [1.0,2.3] |  |
|  | Ivermectin | 33 | 22.6 (18.5) | [16.0,29.1] | 19.6 (7.0,35.9) | 0.0,63.3 | 13.7 [8.8,21.1] |  |

(a) p-value obtained from linear model where mean skin microfilarial density at 1, 6, 12 or 18 months was the outcome variable with treatment as explanatory variable controlling for other covariates such as intensity of infection pre-treatment and sex.
p = <0.0001 for Iol <20, 20-<50, ≥80 for the interaction of Iol category and treatment across SmfD at Month 1, 6, 12, 18, obtained from linear model where mean skin microfilarial density at 1, 6, 12 or 18 months was the outcome variable with treatment as explanatory variable controlling for other covariates such as intensity of infection pre-treatment and sex.
SmfD were logarithmic transformed ( $y'=\log(y+1)$ ) before analysis and back transformed to give geometric means and difference of geometric means in the dimension of the raw data.

**Table S4.** SmfD pre-treatment, 1, 6, 12 and 18 months post treatment in Nord-Ituri (DRC) by intensity of infection pre-treatment.

| Pre-Tx Iol (mf/mg) | Treatment Group | N | Raw Mean (SD) | Raw 95% CI | Raw Median (1Q,3Q) | Raw Min., Max. | Geometric Mean (95%CI) | Overall p-value (a) |
| --- | --- | --- | --- | --- | --- | --- | --- | --- |
| Pre-treatment SmfD |  |  |  |  |  |  |  |  |
| 10 - <20 | Moxidectin | 64 | 13.9 (2.9) | [13.2,14.6] | 13.9 (11.6,16.3) | 9.6,19.5 | 13.6 [12.9,14.4] | <0.0001 |
|  | Ivermectin | 33 | 14.1 (2.9) | [13.1,15.1] | 13.2 (12.0,16.4) | 9.9,19.4 | 13.8 [12.8,14.9] |  |
| 20 - <50 | Moxidectin | 145 | 32.9 (8.5) | [31.5,34.3] | 31.9 (25.5,40.2) | 19.8,49.3 | 31.8 [30.4,33.2] |  |
|  | Ivermectin | 53 | 32.5 (8.8) | [30.0,34.9] | 32.3 (23.7,39.9) | 20.2,49.4 | 31.3 [29.0,33.8] |  |
| ≥50 | Moxidectin | 106 | 89.6 (41.6) | [81.6,97.6] | 73.3 (61.3,104.0) | 49.6,299.4 | 82.7 [76.9,89.0] |  |
|  | Ivermectin | 71 | 85.5 (33.3) | [77.6,93.4] | 77.7 (60.7,97.4) | 50.2,238.3 | 80.7 [74.6,87.2] |  |
| Month 1 |  |  |  |  |  |  |  |  |
| 10 - <20 | Moxidectin | 64 | 0.0 (0.1) | [-0.0,0.0] | 0.0 (0.0,0.0) | 0.0,0.6 | 0.0 [-0.0,0.0] | 0.014 |
|  | Ivermectin | 33 | 1.6 (3.2) | [0.5,2.8] | 0.2 (0.0,1.1) | 0.0,11.5 | 0.8 [0.3,1.3] |  |
| 20 - <50 | Moxidectin | 145 | 0.1 (0.4) | [0.0,0.2] | 0.0 (0.0,0.1) | 0.0,3.4 | 0.1 [0.0,0.1] |  |
|  | Ivermectin | 53 | 2.1 (5.8) | [0.5,3.7] | 0.1 (0.0,1.5) | 0.0,34.6 | 0.8 [0.4,1.2] |  |
| ≥50 | Moxidectin | 105 | 0.4 (1.2) | [0.2,0.7] | 0.0 (0.0,0.3) | 0.0,8.4 | 0.2 [0.1,0.4] |  |
|  | Ivermectin | 71 | 4.7 (12.5) | [1.7,7.6] | 0.2 (0.0,1.7) | 0.0,51.9 | 1.1 [0.6,1.7] |  |
| Month 6 |  |  |  |  |  |  |  |  |
| 10 - <20 | Moxidectin | 64 | 0.0 (0.0) | [-0.0,0.0] | 0.0 (0.0,0.0) | 0.0,0.1 | 0.0 [-0.0,0.0] | <0.0001 |
|  | Ivermectin | 33 | 1.9 (4.0) | [0.5,3.3] | 0.5 (0.0,1.3) | 0.0,21.2 | 0.9 [0.5,1.6] |  |
| 20 - <50 | Moxidectin | 142 | 0.0 (0.1) | [0.0,0.0] | 0.0 (0.0,0.0) | 0.0,0.4 | 0.0 [0.0,0.0] |  |
|  | Ivermectin | 53 | 2.9 (4.1) | [1.8,4.1] | 1.6 (0.3,3.9) | 0.0,21.6 | 1.8 [1.2,2.5] |  |
| ≥50 | Moxidectin | 106 | 0.0 (0.1) | [0.0,0.1] | 0.0 (0.0,0.0) | 0.0,1.2 | 0.0 [0.0,0.1] |  |
|  | Ivermectin | 71 | 8.2 (9.2) | [6.1,10.4] | 4.3 (1.7,11.7) | 0.0,43.1 | 4.8 [3.6,6.4] |  |
| Month 12 |  |  |  |  |  |  |  |  |
| 10 - <20 | Moxidectin | 62 | 0.4 (0.7) | [0.2,0.6] | 0.0 (0.0,0.4) | 0.0,3.6 | 0.3 [0.2,0.4] | <0.0001 |
|  | Ivermectin | 33 | 5.9 (10.8) | [2.1,9.7] | 2.3 (0.5,6.5) | 0.0,57.5 | 2.7 [1.5,4.4] |  |
| 20 - <50 | Moxidectin | 140 | 1.2 (2.5) | [0.8,1.7] | 0.2 (0.0,0.9) | 0.0,14.9 | 0.6 [0.5,0.8] |  |
|  | Ivermectin | 53 | 8.5 (7.9) | [6.4,10.7] | 7.1 (3.0,10.2) | 0.0,35.5 | 5.8 [4.3,7.7] |  |
| ≥50 | Moxidectin | 106 | 2.4 (3.9) | [1.6,3.1] | 0.7 (0.1,3.0) | 0.0,20.1 | 1.2 [0.9,1.6] |  |
|  | Ivermectin | 70 | 22.3 (17.0) | [18.3,26.4] | 17.3 (9.3,34.7) | 0.0,76.5 | 16.1 [12.8,20.1] |  |
| Month 18 |  |  |  |  |  |  |  |  |
| 10 - <20 | Moxidectin | 59 | 2.0 (2.5) | [1.3,2.6] | 0.7 (0.1,3.0) | 0.0,10.3 | 1.2 [0.8,1.7] | <0.0001 |
|  | Ivermectin | 28 | 8.5 (10.2) | [4.5,12.5] | 5.5 (2.1,9.3) | 0.0,41.2 | 5.1 [3.2,7.8] |  |
| 20 - <50 | Moxidectin | 125 | 4.9 (9.1) | [3.3,6.5] | 1.6 (0.3,5.3) | 0.0,63.2 | 2.2 [1.7,2.8] |  |
|  | Ivermectin | 46 | 14.5 (13.5) | [10.5,18.5] | 9.8 (7.0,18.7) | 0.1,57.4 | 9.8 [7.3,13.2] |  |
| ≥50 | Moxidectin | 99 | 11.3 (14.4) | [8.4,14.1] | 5.5 (0.9,18.4) | 0.0,70.5 | 5.3 [4.0,7.0] |  |
|  | Ivermectin | 68 | 35.1 (24.3) | [29.2,41.0] | 30.1 (15.8,50.1) | 0.5,101.3 | 26.4 [21.3,32.6] |  |

(a) p-value obtained from linear model where mean skin microfilarial density at 1, 6, 12 or 18 months was the outcome variable with treatment as explaining variable controlling for other covariates such as intensity of infection pre-treatment and sex.
p = <0.0001 for Iol <20, 20-<50, ≥80 for the interaction of Iol category and treatment across SmfD at Month 1, 6, 12, 18, obtained from linear model where mean skin microfilarial density at 1, 6, 12 or 18 months was the outcome variable with treatment as explanatory variable controlling for other covariates such as intensity of infection pre-treatment and sex.
SmfD were logarithmic transformed ( $y'=\log(y+1)$ ) before analysis and back transformed to give geometric means and difference of geometric means in the dimension of the raw data.

**Table S5.** SmfD pre-treatment, 1, 6, 12 and 18 months post treatment in Lofa County, Liberia, by intensity of infection pre-treatment.

| Pre-Tx Iol (mf/mg) | Treatment Group | N | Raw Mean (SD) | Raw 95% CI | Raw Median (1Q,3Q) | Raw Min., Max. | Geometric Mean (95%CI) | Overall p-value (a) |
| --- | --- | --- | --- | --- | --- | --- | --- | --- |
| Pre-treatment SmfD |  |  |  |  |  |  |  |  |
| 10 - <20 | Moxidectin | 83 | 13.9 (2.8) | [13.3,14.5] | 13.1 (11.8,16.4) | 9.2,19.4 | 13.7 [13.1,14.3] | <0.0001 |
|  | Ivermectin | 42 | 13.5 (2.7) | [12.7,14.3] | 13.3 (10.9,15.3) | 9.7,18.8 | 13.3 [12.5,14.1] |  |
| 20 - <50 | Moxidectin | 84 | 31.9 (7.8) | [30.2,33.6] | 30.7 (24.9,38.5) | 19.5,47.3 | 31.0 [29.4,32.7] |  |
|  | Ivermectin | 39 | 30.2 (7.7) | [27.7,32.7] | 28.0 (23.9,33.4) | 19.7,48.6 | 29.4 [27.1,31.8] |  |
| ≥50 | Moxidectin | 33 | 70.1 (24.2) | [61.5,78.7] | 63.4 (54.9,73.8) | 50.3,157.0 | 67.2 [60.9,74.1] |  |
|  | Ivermectin | 18 | 64.1 (12.2) | [58.0,70.2] | 61.3 (54.6,69.1) | 51.2,90.4 | 63.1 [57.8,69.0] |  |
| Month 1 |  |  |  |  |  |  |  |  |
| 10 - <20 | Moxidectin | 83 | 0.0 (0.1) | [-0.0,0.0] | 0.0 (0.0,0.0) | 0.0,0.5 | 0.0 [-0.0,0.0] | 0.053 |
|  | Ivermectin | 42 | 0.7 (1.8) | [0.1,1.3] | 0.0 (0.0,0.4) | 0.0,9.8 | 0.3 [0.1,0.6] |  |
| 20 - <50 | Moxidectin | 84 | 0.0 (0.1) | [0.0,0.0] | 0.0 (0.0,0.0) | 0.0,0.4 | 0.0 [0.0,0.0] |  |
|  | Ivermectin | 39 | 0.7 (1.3) | [0.2,1.1] | 0.0 (0.0,0.8) | 0.0,6.0 | 0.4 [0.2,0.7] |  |
| ≥50 | Moxidectin | 33 | 0.1 (0.2) | [-0.0,0.1] | 0.0 (0.0,0.0) | 0.0,1.0 | 0.0 [0.0,0.1] |  |
|  | Ivermectin | 18 | 4.3 (12.6) | [-2.0,10.5] | 0.1 (0.0,1.7) | 0.0,53.6 | 1.0 [0.1,2.5] |  |
| Month 6 |  |  |  |  |  |  |  |  |
| 10 - <20 | Moxidectin | 80 | 0.0 (0.0) | [-0.0,0.0] | 0.0 (0.0,0.0) | 0.0,0.3 | 0.0 [-0.0,0.0] | <0.0001 |
|  | Ivermectin | 42 | 0.9 (1.0) | [0.6,1.2] | 0.6 (0.2,1.2) | 0.0,4.0 | 0.7 [0.5,1.0] |  |
| 20 - <50 | Moxidectin | 83 | 0.0 (0.2) | [-0.0,0.1] | 0.0 (0.0,0.0) | 0.0,1.1 | 0.0 [-0.0,0.0] |  |
|  | Ivermectin | 39 | 1.6 (2.6) | [0.8,2.5] | 0.9 (0.3,1.7) | 0.0,15.3 | 1.1 [0.7,1.5] |  |
| ≥50 | Moxidectin | 33 | 0.1 (0.2) | [-0.0,0.1] | 0.0 (0.0,0.0) | 0.0,1.4 | 0.0 [-0.0,0.1] |  |
|  | Ivermectin | 18 | 3.9 (5.2) | [1.3,6.5] | 1.9 (0.5,5.2) | 0.0,21.5 | 2.3 [1.1,4.1] |  |
| Month 12 |  |  |  |  |  |  |  |  |
| 10 - <20 | Moxidectin | 77 | 0.5 (1.3) | [0.2,0.8] | 0.0 (0.0,0.2) | 0.0,8.3 | 0.2 [0.1,0.4] | <0.0001 |
|  | Ivermectin | 41 | 2.4 (2.7) | [1.6,3.3] | 1.4 (0.8,2.9) | 0.0,10.3 | 1.7 [1.1,2.3] |  |
| 20 - <50 | Moxidectin | 78 | 1.1 (2.9) | [0.5,1.8] | 0.1 (0.0,0.8) | 0.0,18.2 | 0.5 [0.3,0.8] |  |
|  | Ivermectin | 36 | 4.7 (6.2) | [2.6,6.8] | 3.0 (1.1,5.4) | 0.0,34.0 | 3.0 [2.0,4.2] |  |
| ≥50 | Moxidectin | 32 | 1.8 (3.8) | [0.5,3.2] | 0.2 (0.0,1.5) | 0.0,15.8 | 0.8 [0.3,1.4] |  |
|  | Ivermectin | 16 | 12.0 (11.1) | [6.1,17.9] | 7.5 (5.4,17.6) | 0.6,43.8 | 8.5 [5.0,13.9] |  |
| Month 18 |  |  |  |  |  |  |  |  |
| 10 - <20 | Moxidectin | 75 | 1.1 (2.2) | [0.6,1.6] | 0.2 (0.0,1.0) | 0.0,14.9 | 0.6 [0.4,0.9] | <0.0001 |
|  | Ivermectin | 40 | 3.5 (3.3) | [2.4,4.5] | 2.1 (1.0,5.1) | 0.0,13.0 | 2.5 [1.7,3.4] |  |
| 20 - <50 | Moxidectin | 75 | 2.0 (3.7) | [1.2,2.9] | 0.3 (0.0,2.0) | 0.0,16.0 | 0.9 [0.6,1.4] |  |
|  | Ivermectin | 37 | 7.2 (9.3) | [4.1,10.3] | 3.4 (2.2,7.7) | 0.0,40.9 | 3.9 [2.5,6.0] |  |
| ≥50 | Moxidectin | 33 | 4.6 (7.1) | [2.1,7.1] | 1.6 (0.0,5.7) | 0.0,27.1 | 2.0 [1.0,3.4] |  |
|  | Ivermectin | 16 | 15.6 (16.5) | [6.8,24.4] | 11.5 (3.9,22.7) | 1.7,66.4 | 10.1 [5.7,17.3] |  |

(a) p-value obtained from linear model where mean skin microfilarial density at 1, 6, 12 or 18 months was the outcome variable with treatment as explaining variable controlling for other covariates such as intensity of infection pre-treatment and sex.
p = <0.0001 for Iol <20, 20-<50, ≥80 for the interaction of Iol category and treatment across SmfD at Month 1, 6, 12, 18, obtained from linear model where mean skin microfilarial density at 1, 6, 12 or 18 months was the outcome variable with treatment as explanatory variable controlling for other covariates such as intensity of infection pre-treatment and sex.
SmfD were logarithmic transformed ( $y'=\log(y+1)$ ) before analysis and back transformed to give geometric means and difference of geometric means in the dimension of the raw data.

**Table S6.** SmfD pre-treatment, 1, 6, 12 and 18 months post treatment in Nkwanta district, Ghana, by intensity of infection pre-treatment.

| Pre-Tx Iol | Treatment Group | N | Raw Mean (SD) | Raw 95% CI | Raw Median (1Q,3Q) | Raw Min., Max. | Geometric Mean (95%CI) | Overall p-value (a) |
| --- | --- | --- | --- | --- | --- | --- | --- | --- |
| Pre-treatment SmfD |  |  |  |  |  |  |  |  |
| 10 - <20 | Moxidectin | 42 | 14.7 (2.8) | [13.9,15.6] | 14.5 (12.3,17.6) | 10.5,19.0 | 14.5 [13.6,15.4] | <0.0001 |
|  | Ivermectin | 24 | 14.5 (2.6) | [13.4,15.6] | 14.5 (13.1,17.1) | 10.0,18.6 | 14.3 [13.2,15.5] |  |
| 20 - <50 | Moxidectin | 76 | 32.5 (9.5) | [30.3,34.6] | 31.9 (23.2,40.5) | 19.6,49.2 | 31.2 [29.1,33.3] |  |
|  | Ivermectin | 38 | 32.1 (8.3) | [29.4,34.8] | 32.1 (24.6,37.8) | 20.4,48.1 | 31.1 [28.5,33.9] |  |
| ≥50 | Moxidectin | 40 | 77.8 (23.3) | [70.4,85.3] | 75.0 (60.5,90.7) | 49.8,160.6 | 75.0 [68.7,81.8] |  |
|  | Ivermectin | 21 | 73.8 (19.4) | [65.0,82.6] | 71.4 (58.5,82.4) | 49.6,124.0 | 71.6 [63.9,80.2] |  |
| Month 1 |  |  |  |  |  |  |  |  |
| 10 - <20 | Moxidectin | 42 | 0.0 (0.0) | [-0.0,0.0] | 0.0 (0.0,0.0) | 0.0,0.2 | 0.0 [-0.0,0.0] | 0.448 |
|  | Ivermectin | 24 | 1.5 (3.7) | [-0.0,3.1] | 0.2 (0.0,0.6) | 0.0,15.9 | 0.6 [0.2,1.3] |  |
| 20 - <50 | Moxidectin | 75 | 0.0 (0.0) | [0.0,0.0] | 0.0 (0.0,0.0) | 0.0,0.2 | 0.0 [0.0,0.0] |  |
|  | Ivermectin | 38 | 2.3 (8.5) | [-0.5,5.1] | 0.0 (0.0,0.5) | 0.0,51.3 | 0.6 [0.2,1.1] |  |
| ≥50 | Moxidectin | 40 | 0.1 (0.2) | [0.0,0.1] | 0.0 (0.0,0.1) | 0.0,1.1 | 0.1 [0.0,0.1] |  |
|  | Ivermectin | 21 | 2.9 (7.7) | [-0.6,6.4] | 0.2 (0.0,2.2) | 0.0,34.8 | 0.9 [0.2,2.0] |  |
| Month 6 |  |  |  |  |  |  |  |  |
| 10 - <20 | Moxidectin | 42 | 0.0 (0.1) | [0.0,0.0] | 0.0 (0.0,0.0) | 0.0,0.3 | 0.0 [0.0,0.0] | 0.105 |
|  | Ivermectin | 24 | 2.5 (3.8) | [0.9,4.1] | 1.1 (0.5,3.0) | 0.2,17.1 | 1.6 [0.9,2.5] |  |
| 20 - <50 | Moxidectin | 76 | 0.0 (0.1) | [0.0,0.1] | 0.0 (0.0,0.0) | 0.0,0.9 | 0.0 [0.0,0.1] |  |
|  | Ivermectin | 38 | 3.4 (5.2) | [1.7,5.1] | 1.8 (0.8,4.8) | 0.0,31.0 | 2.1 [1.4,3.0] |  |
| ≥50 | Moxidectin | 38 | 0.0 (0.1) | [0.0,0.1] | 0.0 (0.0,0.0) | 0.0,0.4 | 0.0 [0.0,0.1] |  |
|  | Ivermectin | 21 | 5.2 (7.0) | [2.1,8.4] | 2.1 (1.6,6.1) | 0.2,29.7 | 3.2 [1.8,5.2] |  |
| Month 12 |  |  |  |  |  |  |  |  |
| 10 - <20 | Moxidectin | 41 | 0.8 (1.6) | [0.3,1.3] | 0.1 (0.0,0.8) | 0.0,8.3 | 0.5 [0.3,0.8] | <0.0001 |
|  | Ivermectin | 23 | 5.5 (5.2) | [3.2,7.7] | 4.4 (1.6,6.5) | 0.0,22.9 | 3.9 [2.4,5.9] |  |
| 20 - <50 | Moxidectin | 75 | 2.5 (4.2) | [1.6,3.5] | 0.6 (0.0,3.4) | 0.0,18.4 | 1.3 [0.8,1.8] |  |
|  | Ivermectin | 38 | 7.8 (8.4) | [5.0,10.6] | 5.6 (2.3,11.3) | 0.0,40.3 | 4.9 [3.4,7.1] |  |
| ≥50 | Moxidectin | 38 | 5.2 (6.8) | [2.9,7.4] | 2.2 (0.1,6.2) | 0.0,27.9 | 2.5 [1.4,4.0] |  |
|  | Ivermectin | 21 | 23.0 (15.7) | [15.8,30.1] | 18.0 (8.1,34.2) | 0.5,49.2 | 17.1 [11.2,26.0] |  |
| Month 18 |  |  |  |  |  |  |  |  |
| 10 - <20 | Moxidectin | 32 | 2.8 (4.2) | [1.3,4.4] | 1.0 (0.0,3.2) | 0.0,18.9 | 1.5 [0.8,2.4] | <0.0001 |
|  | Ivermectin | 15 | 9.6 (7.5) | [5.5,13.8] | 8.2 (3.5,13.1) | 0.0,29.8 | 7.0 [4.0,11.9] |  |
| 20 - <50 | Moxidectin | 49 | 7.9 (11.1) | [4.7,11.1] | 3.1 (1.5,8.2) | 0.0,47.5 | 4.0 [2.7,5.7] |  |
|  | Ivermectin | 23 | 14.6 (10.1) | [10.2,18.9] | 14.1 (6.2,20.2) | 0.2,41.7 | 10.9 [7.3,16.2] |  |
| ≥50 | Moxidectin | 16 | 13.6 (19.2) | [3.4,23.8] | 3.3 (0.9,25.1) | 0.0,70.0 | 5.2 [1.9,12.2] |  |
|  | Ivermectin | 12 | 33.4 (16.5) | [22.9,43.9] | 32.1 (22.9,42.2) | 4.7,61.7 | 28.8 [18.8,43.7] |  |

(a) p-value for the difference in SmfD at the indicated month between Iol categories, obtained from linear model where mean skin microfilarial density at 1, 6, 12 or 18 months was the outcome variable with treatment as explanatory variable controlling for other covariates such as intensity of infection pre-treatment and sex.
p = <0.0001 for Iol <20, 20-<50, ≥50 for the interaction of Iol category and treatment across SmfD at Month 1, 6, 12, 18, obtained from linear model where mean skin microfilarial density at 1, 6, 12 or 18 months was the outcome variable with treatment as explanatory variable controlling for other covariates such as intensity of infection pre-treatment and sex.
SmfD were logarithmic transformed ( $y'=\log(y+1)$ ) before analysis and back transformed to give geometric means and difference of geometric means in the dimension of the raw data.

**Table S7.** Adjusted arithmetic and geometric means and mean differences in SmfD 1, 6, 12 and 18 months post treatment by study area.

| Study area<br>Time post treatment | Treatment<br>Group | Adjusted<br>Mean<br>(95%CI) | Difference of<br>adj. means<br>(95%CI) | Adjusted<br>Geometric Mean<br>(95%CI) | Difference of<br>Adj geometric<br>Mean (95%CI) | Percent<br>Diffe-<br>rence | p-value° |
| --- | --- | --- | --- | --- | --- | --- | --- |
| **Nord Kivu** |  |  |  |  |  |  |  |
| Month 1 | Moxidectin | 0.0 [-0.1;0.1] |  | 0.0 [-0.1;0.1] |  |  |  |
|  | Ivermectin | 0.6 [0.5;0.7] | 0.5 [0.4;0.7] | 0.7 [0.6;0.9] | 0.7 (0.6;0.8) | 98.57 | <0.0001 |
| Month 6 | Moxidectin | -0.0 [-0.1;0.1] |  | -0.0 [-0.1;0.1] |  |  |  |
|  | Ivermectin | 0.9 [0.8;1.0] | 0.9 [0.8;1.0] | 1.4 [1.2;1.7] | 1.4 (1.3;1.6) | 100.42 | <0.0001 |
| Month 12 | Moxidectin | 0.2 [0.1;0.3] |  | 0.2 [0.2;0.3] |  |  |  |
|  | Ivermectin | 1.5 [1.4;1.6] | 1.3 [1.1;1.4] | 3.4 [3.0;3.9] | 3.2 (2.8;3.5) | 92.90 | <0.0001 |
| Month 18 | Moxidectin | 0.6 [0.5;0.6] |  | 0.8 [0.6;0.9] |  |  |  |
|  | Ivermectin | 1.8 [1.6;1.9] | 1.2 [1.1;1.3] | 4.8 [4.2;5.4] | 4.0 (3.5;4.5) | 84.12 | <0.0001 |
| **Nord Ituri** |  |  |  |  |  |  |  |
| Month 1 | Moxidectin | 0.1 [0.0;0.2] |  | 0.1 [0.0;0.2] |  |  |  |
|  | Ivermectin | 0.6 [0.4;0.7] | 0.5 [0.3;0.6] | 0.8 [0.6;1.0] | 0.6 (0.5;0.8) | 83.70 | <0.0001 |
| Month 6 | Moxidectin | 0.0 [-0.1;0.1] |  | 0.0 [-0.1;0.1] |  |  |  |
|  | Ivermectin | 1.2 [1.1;1.3] | 1.2 [1.0;1.3] | 2.4 [2.0;2.8] | 2.3 (2.0;2.7) | 98.76 | <0.0001 |
| Month 12 | Moxidectin | 0.6 [0.5;0.6] |  | 0.7 [0.6;0.9] |  |  |  |
|  | Ivermectin | 2.1 [2.0;2.3] | 1.6 [1.4;1.7] | 7.5 [6.5;8.7] | 6.8 (5.9;7.7) | 90.14 | <0.0001 |
| Month 18 | Moxidectin | 1.3 [1.2;1.4] |  | 2.8 [2.5;3.2] |  |  |  |
|  | Ivermectin | 2.6 [2.5;2.8] | 1.3 [1.1;1.4] | 12.8 [11.1;14.8] | 10.0 (8.6;11.6) | 77.97 | <0.0001 |
| **Lofa County** |  |  |  |  |  |  |  |
| Month 1 | Moxidectin | 0.0 [-0.1;0.1] |  | 0.0 [-0.1;0.1] |  |  |  |
|  | Ivermectin | 0.5 [0.4;0.6] | 0.5 [0.3;0.6] | 0.6 [0.4;0.9] | 0.6 (0.5;0.7) | 97.07 | <0.0001 |
| Month 6 | Moxidectin | 0.0 [-0.1;0.1] |  | 0.0 [-0.1;0.1] |  |  |  |
|  | Ivermectin | 0.8 [0.7;1.0] | 0.8 [0.7;1.0] | 1.3 [1.0;1.6] | 1.3 (1.1;1.5) | 98.75 | <0.0001 |
| Month 12 | Moxidectin | 0.4 [0.3;0.5] |  | 0.4 [0.3;0.6] |  |  |  |
|  | Ivermectin | 1.5 [1.3;1.6] | 1.1 [0.9;1.3] | 3.3 [2.8;3.9] | 2.9 (2.5;3.3) | 86.75 | <0.0001 |
| Month 18 | Moxidectin | 0.7 [0.6;0.8] |  | 1.0 [0.8;1.2] |  |  |  |
|  | Ivermectin | 1.7 [1.6;1.8] | 1.0 [0.9;1.2] | 4.4 [3.8;5.2] | 3.5 (3.0;4.0) | 78.23 | <0.0001 |
| **Nkwanta N** |  |  |  |  |  |  |  |
| Month 1 | Moxidectin | -0.1 [-0.2;0.0] |  | -0.1 [-0.2;0.0] |  |  |  |
|  | Ivermectin | 0.5 [0.4;0.7] | 0.6 [0.4;0.8] | 0.7 [0.4;1.0] | 0.8 (0.6;1.0) | 110.69 | <0.0001 |
| Month 6 | Moxidectin | -0.1 [-0.2;0.1] |  | -0.1 [-0.2;0.1] |  |  |  |
|  | Ivermectin | 1.2 [1.0;1.3] | 1.2 [1.0;1.5] | 2.2 [1.7;2.8] | 2.3 (1.9;2.8) | 103.21 | <0.0001 |
| Month 12 | Moxidectin | 0.7 [0.6;0.8] |  | 1.0 [0.8;1.3] |  |  |  |
|  | Ivermectin | 2.0 [1.9;2.2] | 1.3 [1.1;1.5] | 6.6 [5.4;8.0] | 5.6 (4.6;6.7) | 84.48 | <0.0001 |
| Month 18 | Moxidectin | 1.3 [1.2;1.5] |  | 2.9 [2.3;3.5] |  |  |  |
|  | Ivermectin | 2.6 [2.4;2.8] | 1.3 [1.0;1.5] | 13.1 [10.5;16.2] | 10.2 (8.2;12.8) | 78.15 | <0.0001 |

p-value for the difference in SmfD after moxidectin and ivermectin at each follow up time point obtained by marginal (least square means) means pairwise comparisons from the mixed effect model per study area where microfilarial density at 1, 6, 12 and 18 months was the outcome variable with treatment, follow up time point (1, 6, 12, 18 months), and interaction between treatment and follow up time point and interaction between treatment and intensity of infection at baseline as fixed effect, controlling for other covariates such as baseline density, baseline intensity of infection and sex.
>100% difference due to low SmfD values in the moxidectin treatment arm that are log-transformed to calculate the geometric means.

**Table S8.** Adjusted SmfD means and mean differences 12 months post-treatment by pre-treatment IoI.

| Study area<br>Screening intensity of<br>Infection | Treatment<br>Group | Adjusted<br>Mean<br>(95%CI) | Difference of<br>adj. means<br>(95%CI) | Adjusted<br>Geometric<br>Mean<br>(95%CI) | Difference of<br>Adj geometric<br>Mean (95%CI) | Percent<br>Difference | p-value° |
| --- | --- | --- | --- | --- | --- | --- | --- |
| **Across all** |  |  |  |  |  |  |  |
| 10 - <20 mf/mg | Moxidectin | 0.5 [0.3;0.7] |  | 0.6 [0.3;0.9] |  |  |  |
|  | Ivermectin | 1.2 [1.0;1.4] | 0.7 [0.6;0.8] | 2.3 [1.7;3.0] | 1.7 (1.4;2.1) | 74.10 | <0.0001 |
| 20 - <50 mf/mg | Moxidectin | 0.3 [0.2;0.5] |  | 0.4 [0.2;0.6] |  |  |  |
|  | Ivermectin | 1.3 [1.1;1.4] | 0.9 [0.8;1.0] | 2.6 [2.0;3.2] | 2.2 (1.8;2.6) | 84.21 | <0.0001 |
| 50 - <80 mf/mg | Moxidectin | 0.3 [0.1;0.5] |  | 0.4 [0.1;0.7] |  |  |  |
|  | Ivermectin | 1.5 [1.4;1.7] | 1.2 [1.1;1.4] | 3.7 [2.9;4.7] | 3.3 (2.7;4.1) | 89.98 | <0.0001 |
| ≥80 mf/mg | Moxidectin | 0.2 [-0.0;0.5] |  | 0.3 [0.0;0.6] |  |  |  |
|  | Ivermectin | 1.9 [1.7;2.2] | 1.7 [1.5;1.9] | 5.7 [4.2;7.6] | 5.5 (4.2;7.0) | 95.38 | <0.0001 |
| **Nord Kivu** |  |  |  |  |  |  |  |
| 10 - <20 mf/mg | Moxidectin | 0.5 [0.3;0.6] |  | 0.6 [0.3;0.9] |  |  |  |
|  | Ivermectin | 1.0 [0.8;1.2] | 0.6 [0.4;0.7] | 1.8 [1.3;2.4] | 1.2 (1.0;1.5) | 68.15 | <0.0001 |
| 20 - <50 mf/mg | Moxidectin | 0.2 [0.1;0.3] |  | 0.2 [0.1;0.3] |  |  |  |
|  | Ivermectin | 1.1 [1.0;1.2] | 0.9 [0.7;1.1] | 2.0 [1.6;2.4] | 1.8 (1.5;2.1) | 89.62 | <0.0001 |
| ≥50 mf/mg | Moxidectin | -0.1 [-0.2;0.1] |  | 0.0 [-0.2;0.2] |  |  |  |
|  | Ivermectin | 1.4 [1.2;1.6] | 1.4 [1.2;1.6] | 3.0 [2.2;3.9] | 3.0 (2.5;3.8) | 101.63 | <0.0001 |
| **Nord Ituri** |  |  |  |  |  |  |  |
| 10 - <20 mf/mg | Moxidectin | 0.6 [0.4;0.9] |  | 0.9 [0.5;1.4] |  |  |  |
|  | Ivermectin | 1.6 [1.3;1.8] | 0.9 [0.7;1.2] | 3.7 [2.6;5.2] | 2.8 (2.1;3.8) | 75.73 | <0.0001 |
| 20 - <50 mf/mg | Moxidectin | 0.5 [0.4;0.6] |  | 0.6 [0.5;0.8] |  |  |  |
|  | Ivermectin | 1.6 [1.4;1.7] | 1.1 [0.9;1.3] | 3.8 [3.1;4.7] | 3.2 (2.7;3.9) | 83.90 | <0.0001 |
| ≥50 mf/mg | Moxidectin | 0.4 [0.2;0.6] |  | 0.5 [0.3;0.8] |  |  |  |
|  | Ivermectin | 1.8 [1.6;2.0] | 1.4 [1.2;1.6] | 4.9 [3.9;6.2] | 4.4 (3.7;5.4) | 89.91 | <0.0001 |
| **Lofa County** |  |  |  |  |  |  |  |
| 10 - <20 mf/mg | Moxidectin | 0.2 [0.1;0.4] |  | 0.2 [0.1;0.5] |  |  |  |
|  | Ivermectin | 0.8 [0.6;1.0] | 0.6 [0.5;0.8] | 1.3 [0.9;1.8] | 1.1 (0.9;1.3) | 81.16 | <0.0001 |
| 20 - <50 mf/mg | Moxidectin | 0.2 [0.1;0.3] |  | 0.3 [0.1;0.4] |  |  |  |
|  | Ivermectin | 1.0 [0.9;1.1] | 0.8 [0.6;0.9] | 1.7 [1.3;2.1] | 1.5 (1.2;1.7) | 85.29 | <0.0001 |
| ≥50 mf/mg | Moxidectin | 0.4 [0.1;0.6] |  | 0.4 [0.1;0.9] |  |  |  |
|  | Ivermectin | 1.5 [1.2;1.8] | 1.2 [0.9;1.4] | 3.6 [2.4;5.1] | 3.1 (2.3;4.2) | 87.85 | <0.0001 |
| **Nkwanta** |  |  |  |  |  |  |  |
| 10 - <20 mf/mg | Moxidectin | 0.4 [0.1;0.7] |  | 0.5 [0.2;1.0] |  |  |  |
|  | Ivermectin | 1.4 [1.1;1.8] | 1.0 [0.7;1.3] | 3.2 [2.1;4.8] | 2.7 (1.9;3.8) | 83.87 | <0.0001 |
| 20 - <50 mf/mg | Moxidectin | 0.5 [0.4;0.6] |  | 0.6 [0.4;0.9] |  |  |  |
|  | Ivermectin | 1.4 [1.3;1.6] | 1.0 [0.7;1.2] | 3.3 [2.5;4.2] | 2.6 (2.1;3.3) | 80.18 | <0.0001 |
| ≥50 mf/mg | Moxidectin | 0.5 [0.2;0.8] |  | 0.7 [0.2;1.3] |  |  |  |
|  | Ivermectin | 1.9 [1.6;2.2] | 1.4 [1.1;1.7] | 5.7 [3.8;8.4] | 5.0 (3.5;7.1) | 88.22 | <0.0001 |

p-value for the difference in SmfD after moxidectin and ivermectin for each lol category obtained by marginal (least square means) means pairwise comparisons from the mixed effect model across and per study area where microfilarial density at 12 months was the outcome variable with treatment, baseline intensity of infection and sex as covariates

**Table S9.** Participants with undetectable SmfD from Month 1 to 6, 12 or 18 by IoI and area.

| Area | Moxidectin |  |  |  |  | Ivermectin |  |  |  |  |
| --- | --- | --- | --- | --- | --- | --- | --- | --- | --- | --- |
| lol | ND | n UD Yes | n UD No | N | % with UD | ND | n UD Yes | n UD No | N | % with UD |
| **All areas** |  |  |  |  |  |  |  |  |  |  |
| UD 1-6 |  |  |  |  |  |  |  |  |  |  |
| Any | 20 | 741 | 217 | 958 | 77.3 | 5 | 35 | 454 | 489 | 7.2 |
| 10-<20 | 6 | 245 | 30 | 275 | 89.1 |  | 18 | 132 | 150 | 12 |
| 20-<50 | 10 | 354 | 92 | 446 | 79.4 | 2 | 14 | 167 | 181 | 7.7 |
| 50-<80 | 3 | 107 | 47 | 154 | 69.5 | 3 | 3 | 102 | 105 | 2.9 |
| ≥80 | 1 | 35 | 48 | 83 | 42.2 |  | 0 | 53 | 53 | 0 |
| UD 1-12 |  |  |  |  |  |  |  |  |  |  |
| Any | 40 | 360 | 578 | 938 | 38.4 | 16 | 7 | 471 | 478 | 1.5 |
| <20 | 14 | 159 | 108 | 267 | 59.6 | 2 | 3 | 145 | 148 | 2 |
| 20-<50 | 20 | 156 | 280 | 436 | 35.8 | 6 | 3 | 174 | 177 | 1.7 |
| 50-<80 | 4 | 32 | 121 | 153 | 20.9 | 7 | 1 | 100 | 101 | 1 |
| 80+ | 2 | 13 | 69 | 82 | 15.9 | 1 | 0 | 52 | 52 | 0 |
| UD 1-18 |  |  |  |  |  |  |  |  |  |  |
| Any | 222 | 164 | 592 | 756 | 21.7 | 111 | 4 | 379 | 383 | 1 |
| <20 | 55 | 76 | 150 | 226 | 33.6 | 33 | 1 | 116 | 117 | 0.9 |
| 20-<50 | 124 | 68 | 264 | 332 | 20.5 | 44 | 2 | 137 | 139 | 1.4 |
| 50-<80 | 27 | 15 | 115 | 130 | 11.5 | 25 | 1 | 82 | 83 | 1.2 |
| 80+ | 16 | 5 | 63 | 68 | 7.4 | 9 | 0 | 44 | 44 | 0 |
| **N Kivu** |  |  |  |  |  |  |  |  |  |  |
| UD 1-6 |  |  |  |  |  |  |  |  |  |  |
| Any | 9 | 246 | 50 | 296 | 83.1 | 3 | 17 | 133 | 150 | 11.3 |
| 10-<20 | 3 | 80 | 9 | 89 | 89.9 |  | 7 | 44 | 51 | 13.7 |
| 20-<50 | 5 | 123 | 23 | 146 | 84.2 | 2 | 8 | 43 | 51 | 15.7 |
| ≥50 | 1 | 43 | 18 | 61 | 70.5 | 1 | 2 | 46 | 48 | 4.2 |
| UD 1-12 |  |  |  |  |  |  |  |  |  |  |
| Any | 14 | 145 | 146 | 291 | 49.8 | 9 | 4 | 143 | 147 | 2.7 |
| <20 | 5 | 60 | 27 | 87 | 69 |  | 2 | 49 | 51 | 3.9 |
| 20-<50 | 7 | 67 | 77 | 144 | 46.5 | 4 | 1 | 49 | 50 | 2 |
| ≥50 | 2 | 18 | 42 | 60 | 30 | 5 | 1 | 45 | 46 | 2.2 |
| UD 1-18 |  |  |  |  |  |  |  |  |  |  |
| Any | 106 | 73 | 126 | 199 | 36.7 | 55 | 3 | 97 | 100 | 3 |
| <20 | 31 | 31 | 30 | 61 | 50.8 | 17 | 1 | 33 | 34 | 2.9 |
| 20-<50 | 65 | 32 | 54 | 86 | 37.2 | 19 | 1 | 33 | 34 | 2.9 |
| ≥50 | 10 | 10 | 42 | 52 | 19.2 | 19 | 1 | 31 | 32 | 3.1 |
| **N Ituri** |  |  |  |  |  |  |  |  |  |  |
| UD 1-6 |  |  |  |  |  |  |  |  |  |  |
| Any | 4 | 205 | 106 | 311 | 65.9 | 0 | 9 | 148 | 157 | 5.7 |
| <20 | 0 | 56 | 8 | 64 | 87.5 |  | 5 | 28 | 33 | 15.2 |
| 20-<50 | 3 | 96 | 46 | 142 | 67.6 |  | 3 | 50 | 53 | 5.7 |
| ≥50 | 1 | 53 | 52 | 105 | 50.5 | 0 | 1 | 70 | 71 | 1.4 |
| UD 1-12 |  |  |  |  |  |  |  |  |  |  |
| Any | 9 | 77 | 230 | 307 | 25.1 | 1 | 1 | 155 | 156 | 0.6 |
| <20 | 2 | 29 | 33 | 62 | 46.8 |  | 1 | 32 | 33 | 3 |
| 20-<50 | 5 | 36 | 104 | 140 | 25.7 |  | 0 | 53 | 53 | 0 |
| ≥50 | 1 | 12 | 93 | 105 | 11.4 | 1 | 0 | 70 | 70 | 0 |
| UD 1-18 |  |  |  |  |  |  |  |  |  |  |
| Any | 33 | 24 | 258 | 282 | 8.5 | 16 | 0 | 141 | 141 | 0 |
| <20 | 5 | 8 | 51 | 59 | 13.6 | 5 | 0 | 28 | 28 | 0 |
| 20-<50 | 20 | 12 | 113 | 125 | 9.6 | 7 | 0 | 46 | 46 | 0 |
| ≥50 | 8 | 4 | 94 | 98 | 4.1 | 4 | 0 | 67 | 67 | 0 |
| **Lofa** |  |  |  |  |  |  |  |  |  |  |
| UD 1-6 |  |  |  |  |  |  |  |  |  |  |
| Any | 4 | 173 | 23 | 196 | 88.3 | 0 | 8 | 91 | 99 | 8.1 |
| <20 | 3 | 76 | 4 | 80 | 95 |  | 6 | 36 | 42 | 14.3 |
| 20-<50 | 1 | 73 | 10 | 83 | 88 |  | 2 | 37 | 39 | 5.1 |
| ≥50 | 0 | 24 | 9 | 33 | 72.7 | 0 | 0 | 18 | 18 | 0 |
| lol | ND | n UD Yes | n UD No | N | % with UD | ND | n UD Yes | n UD No | N | % with UD |
| UD 1-12 |  |  |  |  |  |  |  |  |  |  |
| Any | 13 | 99 | 88 | 187 | 52.9 | 6 | 1 | 92 | 93 | 1.1 |
| <20 | 6 | 52 | 25 | 77 | 67.5 | 1 | 0 | 41 | 41 | 0 |
| 20-<50 | 6 | 36 | 42 | 78 | 46.2 | 3 | 1 | 35 | 36 | 2.8 |
| ≥50 | 1 | 11 | 21 | 32 | 34.4 | 2 | 0 | 16 | 16 | 0 |
| UD 1-18 |  |  |  |  |  |  |  |  |  |  |
| Any | 20 | 57 | 123 | 180 | 31.7 | 7 | 1 | 91 | 92 | 1.1 |
| <20 | 9 | 31 | 43 | 74 | 41.9 | 2 | 0 | 40 | 40 | 0 |
| 20-<50 | 10 | 23 | 51 | 74 | 31.1 | 3 | 1 | 35 | 36 | 2.8 |
| ≥50 | 1 | 3 | 29 | 32 | 9.4 | 2 | 0 | 16 | 16 | 0 |
| Nkwanta |  |  |  |  |  |  |  |  |  |  |
| UD 1-6 |  |  |  |  |  |  |  |  |  |  |
| Any | 3 | 117 | 38 | 155 | 75.5 | 0 | 1 | 82 | 83 | 1.2 |
| <20 | 0 | 33 | 9 | 42 | 78.6 |  | 0 | 24 | 24 | 0 |
| 20-<50 | 1 | 62 | 13 | 75 | 82.7 |  | 1 | 37 | 38 | 2.6 |
| ≥50 | 2 | 22 | 16 | 38 | 57.9 | 0 | 0 | 21 | 21 | 0 |
| UD 1-12 |  |  |  |  |  |  |  |  |  |  |
| Any | 5 | 39 | 114 | 153 | 25.5 | 1 | 1 | 81 | 82 | 1.2 |
| <20 | 1 | 18 | 23 | 41 | 43.9 | 1 | 0 | 23 | 23 | 0 |
| 20-<50 | 2 | 17 | 57 | 74 | 23 |  | 1 | 37 | 38 | 2.6 |
| ≥50 | 2 | 4 | 34 | 38 | 10.5 | 0 | 0 | 21 | 21 | 0 |
| UD 1-18 |  |  |  |  |  |  |  |  |  |  |
| Any | 63 | 10 | 85 | 95 | 10.5 | 33 | 0 | 50 | 50 | 0 |
| <20 | 10 | 6 | 26 | 32 | 18.8 | 9 | 0 | 15 | 15 | 0 |
| 20-<50 | 29 | 1 | 46 | 47 | 2.1 | 15 | 0 | 23 | 23 | 0 |
| ≥50 | 24 | 3 | 13 | 16 | 18.8 | 9 | 0 | 12 | 12 | 0 |

lol: lol category; N: Number of participants included in the analysis; NC: OR not calculated since Odds for ivermectin group 0; ND: number of participants with a missing data point for at least one of the relevant time points and excluded from the analysis; n UD No: number of participants with data points at all relevant timepoints and detectable SmfD included in the analysis; n UD yes: number of participants with data points at all relevant timepoints and undetectable SmfD included in the analysis, OR: Odds ratio; UD 1-6, UD 1-12, UD 1-18: undetectable SmfD at M1 and M6, at M1, M6 and M12, at M1, M6, M12, M18, respectively.

**Fig S1:**
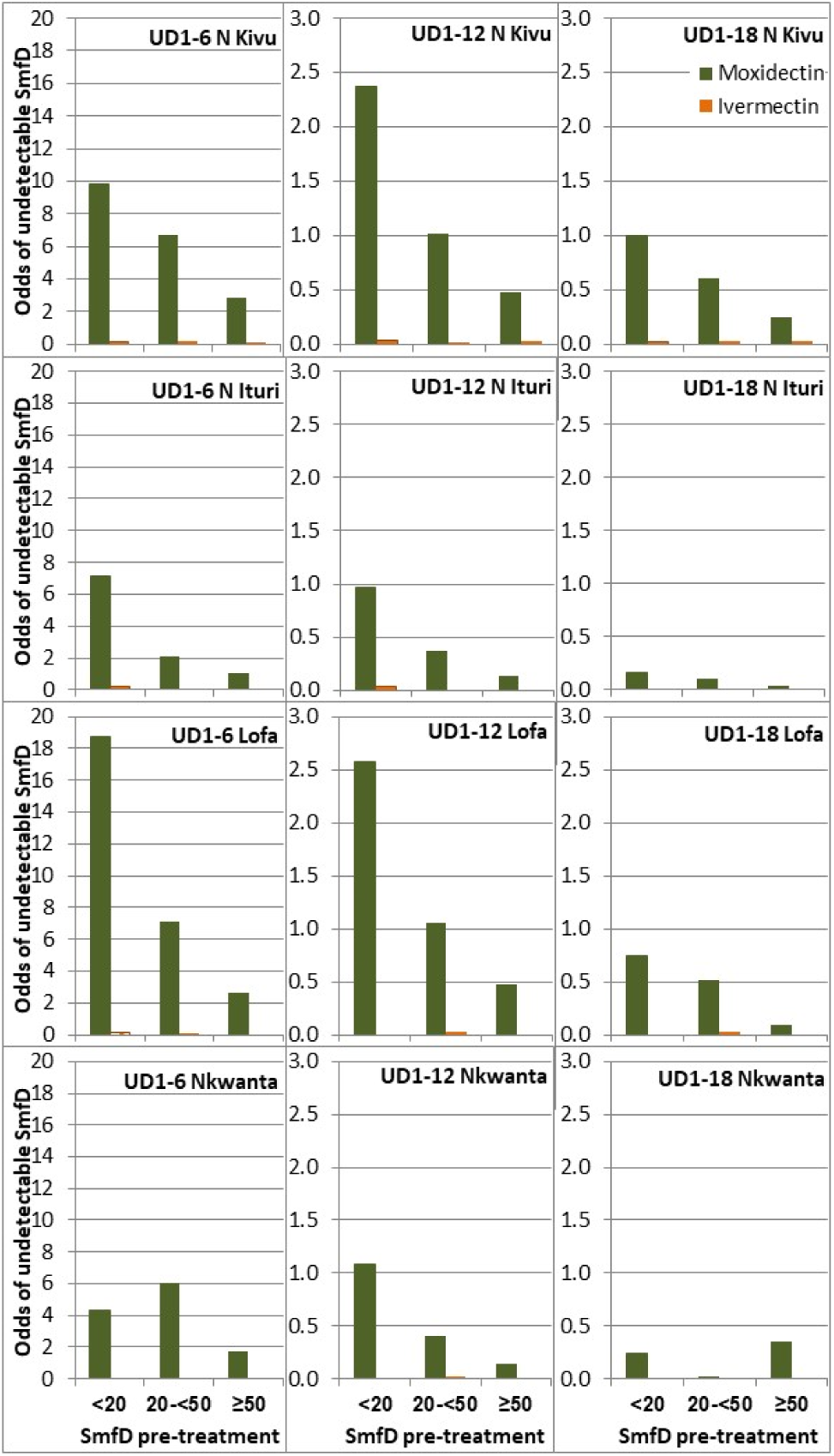
Odds for UD from 1 to 6, 12 or 18 months by IoI and study area. SmfD: skin microfilariae density, UD1-6, UD 1-12, UD 1-18 undetectable levels of SmfD at month 1 and 6, at months 1, 6 and 12, and at month 1, 6, 12 and 18, respectively (for number with follow up data at each relevant time point, see **Error! Reference source not found.**).

**Table S10.** Logistic model derived odds for undetectable levels of skin microfilariae from month 1 sustained to month 6, month 12 or month 18 among participants treated with moxidectin or ivermectin by IoI across and by study area.

| Study area<br>lol | Moxidectin: |  |  |  |  |  |  |  |  | Ivermectin |  |  |  |  |  |  |  |  |
| --- | --- | --- | --- | --- | --- | --- | --- | --- | --- | --- | --- | --- | --- | --- | --- | --- | --- | --- |
|  | M1 and M6 |  |  | M1, M6, and M12 |  |  | M1, M6, M12, and M18 |  |  | M1 and M6 |  |  | M1, M6, and M12 |  |  | M1, M6, M12, and M18 |  |  |
|  | Odds | LL | UL | Odds | LL | UL | Odds | LL | UL | Odds | LL | UL | Odds | LL | UL | Odds | LL | UL |
| **Across all** |  |  |  |  |  |  |  |  |  |  |  |  |  |  |  |  |  |  |
| 10-<20 | 8.48 | 5.12 | 14.05 | 1.54 | 0.92 | 2.59 | 0.41 | 0.18 | 0.93 | 0.13 | 0.07 | 0.23 | 0.02 | 0.00 | 0.05 | 0.01 | 0.00 | 0.04 |
| 20-<50 | 4.33 | 2.88 | 6.51 | 0.62 | 0.38 | 1.02 | 0.24 | 0.11 | 0.54 | 0.08 | 0.04 | 0.16 | 0.02 | 0.00 | 0.05 | 0.01 | 0.00 | 0.06 |
| 50-<80 | 2.59 | 1.60 | 4.20 | 0.27 | 0.15 | 0.50 | 0.10 | 0.04 | 0.27 | 0.03 | 0.01 | 0.10 | 0.01 | 0.00 | 0.07 | 0.01 | 0.00 | 0.08 |
| ≥80 | 0.90 | 0.52 | 1.58 | 0.23 | 0.11 | 0.48 | 0.09 | 0.03 | 0.32 | 0.00 | 0.00 | Infty | 0.00 | 0.00 | Infty | 0.00 | 0.00 | Infty |
| **Nord-Kivu** |  |  |  |  |  |  |  |  |  |  |  |  |  |  |  |  |  |  |
| 10-<20 | 9.84 | 4.85 | 19.97 | 2.37 | 1.48 | 3.79 | 1.00 | 0.60 | 1.66 | 0.15 | 0.07 | 0.34 | 0.04 | 0.01 | 0.16 | 0.03 | 0.00 | 0.21 |
| 20-<50 | 6.70 | 4.11 | 10.90 | 1.02 | 0.72 | 1.45 | 0.60 | 0.39 | 0.94 | 0.18 | 0.09 | 0.40 | 0.02 | 0.00 | 0.14 | 0.03 | 0.00 | 0.22 |
| ≥50 | 2.89 | 1.61 | 5.17 | 0.48 | 0.27 | 0.84 | 0.25 | 0.12 | 0.50 | 0.05 | 0.01 | 0.23 | 0.03 | 0.00 | 0.20 | 0.04 | 0.00 | 0.26 |
| **Nord Ituri** |  |  |  |  |  |  |  |  |  |  |  |  |  |  |  |  |  |  |
| 10-<20 | 7.11 | 3.37 | 15.01 | 0.96 | 0.58 | 1.61 | 0.17 | 0.08 | 0.34 | 0.18 | 0.07 | 0.47 | 0.03 | 0.00 | 0.24 | 0.00 | 0.00 | Infty |
| 20-<50 | 2.12 | 1.48 | 3.04 | 0.37 | 0.26 | 0.52 | 0.11 | 0.06 | 0.20 | 0.06 | 0.02 | 0.20 | 0.00 | 0.00 | Infty | 0.00 | 0.00 | Infty |
| ≥50 | 1.03 | 0.70 | 1.51 | 0.13 | 0.07 | 0.24 | 0.04 | 0.02 | 0.12 | 0.01 | 0.00 | 0.10 | 0.00 | 0.00 | Infty | 0.00 | 0.00 | Infty |
| **Lofa** |  |  |  |  |  |  |  |  |  |  |  |  |  |  |  |  |  |  |
| 10-<20 | 18.71 | 6.81 | 51.41 | 2.57 | 1.81 | 3.66 | 0.75 | 0.46 | 1.22 | 0.16 | 0.07 | 0.39 | 0.00 | 0.00 | Infty | 0.00 | 0.00 | Infty |
| 20-<50 | 7.09 | 3.63 | 13.83 | 1.06 | 0.65 | 1.72 | 0.52 | 0.31 | 0.87 | 0.05 | 0.01 | 0.22 | 0.03 | 0.00 | 0.22 | 0.03 | 0.00 | 0.23 |
| ≥50 | 2.67 | 1.23 | 5.76 | 0.48 | 0.22 | 1.05 | 0.09 | 0.03 | 0.30 | 0.00 | 0.00 | Infty | 0.00 | 0.00 | Infty | 0.00 | 0.00 | Infty |
| **Nkwanta** |  |  |  |  |  |  |  |  |  |  |  |  |  |  |  |  |  |  |
| 10-<20 | 4.32 | 1.92 | 9.73 | 1.08 | 0.61 | 1.90 | 0.24 | 0.09 | 0.64 | 0.00 | 0.00 | Infty | 0.00 | 0.00 | Infty | 0.00 | 0.00 | Infty |
| 20-<50 | 6.04 | 3.06 | 11.92 | 0.40 | 0.22 | 0.74 | 0.03 | 0.00 | 0.21 | 0.03 | 0.00 | 0.23 | 0.03 | 0.00 | 0.21 | 0.00 | 0.00 | Infty |
| ≥50 | 1.73 | 0.85 | 3.54 | 0.14 | 0.05 | 0.43 | 0.35 | 0.09 | 1.35 | 0.00 | 0.00 | Infty | 0.00 | 0.00 | Infty | 0.00 | 0.00 | Infty |

**Table S11.**
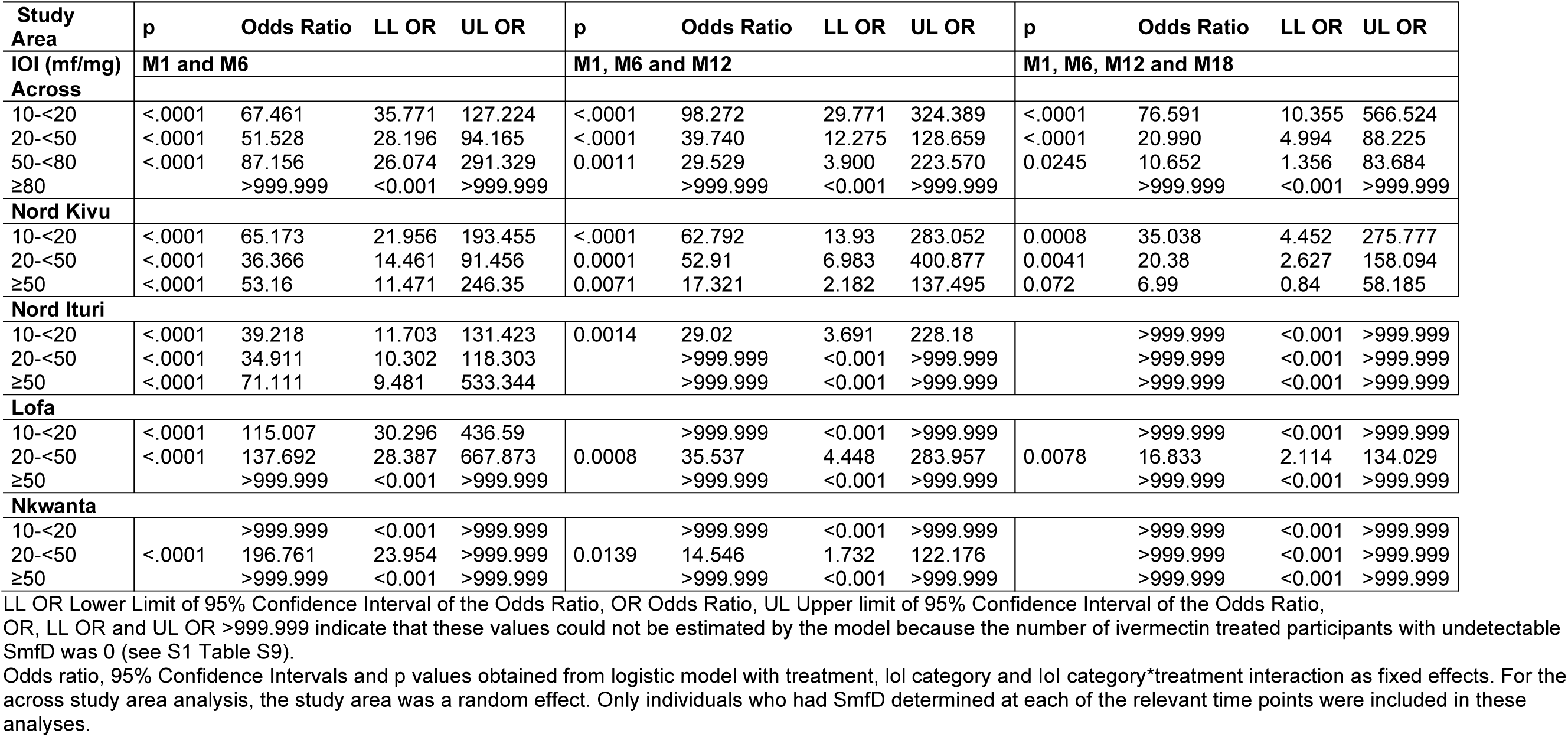
Logistic model derived odds ratios for undetectable levels of skin microfilariae from month 1 sustained to month 6, month 12 or month 18 among participants treated with moxidectin relative to among ivermectin treated participants by IoI by study area.

## Notes

### Clinical Trial

NCT00790998

### Author Declarations

Conduct of this study, including protocol, information documents for potential participants and the participant consent forms, were approved by the Ghana Food and Drugs Authority and the Ghana Health Service Ethics Review Committee, the Liberia Ministry of Health and Social Welfare and the Ethics Committee of the Liberia Institute for Biomedical Research, the Ministere de la Sante Publique of DRC and the Ethics Committee of the Ecole de la Sante Publique Universite de Kinshasa in DRC, and the WHO Ethics Review Committee.

